# Estimating Probabilities of Success of Clinical Trials for Vaccines and Other Anti-Infective Therapeutics

**DOI:** 10.1101/2020.04.09.20059600

**Authors:** Chi Heem Wong, Kien Wei Siah, Andrew W. Lo

## Abstract

A key driver in biopharmaceutical investment decisions is the probability of success of a drug development program. We estimate the probabilities of success (PoS) of clinical trials for vaccines and other anti-infective therapeutics using 43,414 unique triplets of clinical trial, drug, and disease between January 1, 2000, and January 7, 2020, yielding 2,544 vaccine programs and 6,829 non-vaccine programs targeting infectious diseases. The overall estimated PoS for an industry-sponsored vaccine program is 39.6%, and 16.3% for an industry-sponsored anti-infective therapeutic. Among industry-sponsored vaccines programs, only 12 out of 27 disease categories have seen at least one approval, with the most successful being against monkeypox (100%), rotavirus (78.7%), and Japanese encephalitis (67.6%). The three infectious diseases with the highest PoS for industry-sponsored non-vaccine therapeutics are smallpox (100%), CMV (31.8%), and onychomycosis (29.8%). Non-industry-sponsored vaccine and non-vaccine development programs have lower overall PoSs: 6.8% and 8.2%, respectively. Viruses involved in recent outbreaks—MERS, SARS, Ebola, Zika—have had a combined total of only 45 non-vaccine development programs initiated over the past two decades, and no approved therapy to date (Note: our data was obtained just before the COVID-19 outbreak and do not contain information about the programs targeting this disease.) These estimates offer guidance both to biopharma investors as well as to policymakers seeking to identify areas most likely to be undeserved by private-sector engagement and in need of public-sector support.

## Introduction

In this paper, we provide estimates of the historical probabilities of success (PoS) of clinical trials for vaccines and other therapeutic drugs for infectious diseases to inform discussions on the planning and financing of the fight against one of humanity’s oldest foes. This is of particular importance in light of the recent havoc wreaked by the severe acute respiratory syndrome coronavirus 2 (SARS-CoV-2), the virus that causes coronavirus disease (COVID-19).

While the probabilities of success of therapeutic drugs for various disease groups like oncology are well-documented (Abrantes-Metz et al., 2004; DiMasi et al., 2010; Hay et al., 2014; MIT Laboratory for Financial Engineering, 2020; Smietana et al., 2016; Thomas et al., 2016; Wong et al., 2019b, 2019a), relatively little has been published on treatments for infectious diseases and vaccine development despite their importance (Davis et al., 2011; Pronker et al., 2013). Prior studies have focused on narrower subsets relevant to their specific interests and have relied on much more limited data sets. For example, Young et al. (2020) employed 10 to 25 data points per estimated value from the Bill and Melinda Gates Foundation to estimate the PoS of vaccines for neglected diseases, and DiMasi et al. (2020) reported PoS estimates on a per-drug basis using 2,575 trials for diseases of interest to the Gates Foundation. In contrast, we employ a much larger and broader dataset of 16,328 unique clinical trials to estimate the PoS of vaccines and non-vaccine therapeutics targeting 29 different infectious diseases using all available drug/indication pairs—a methodology that has been argued to be more relevant for evaluating drug development R&D risk and productivity (Hay et al., 2014; Thomas et al., 2016; Wong et al., 2019b).

Vaccination is commonly recognized as one of the most cost-effective public health measures for combatting infectious diseases (André, 2002; Ehreth, 2003; Kieny & Girard, 2005; OECD, 2013; Pronker et al., 2013; Rémy et al., 2015). In developed countries, routine prophylactic vaccination and effective treatment options have led to the control or complete elimination of several deadly infectious diseases through individual and herd immunity, preventing millions of deaths and untold suffering each year. This prophylaxis dramatically reduces the burden on the healthcare system and society as a whole. In addition, the deaths, hospitalizations, and treatment costs avoided by these measures have led to significant economic savings (Ehreth, 2003; Rémy et al., 2015; US Department of Health and Human Services, 2017).

As technology continues to advance, one expects that the human species will be better able to cope with these diseases. The fact remains, however, that we still do not have effective treatments or vaccines for many infectious diseases. While the discovery of antibiotics has reduced the mortality rate of bacterial infection, and the development of the smallpox vaccine has led to the eradication of the devastating disease (World Health Organization, 1980), other infectious diseases, such as Acquired Immunodeficiency Syndrome (AIDS) and malaria, still take the lives of tens of millions every year. According to the World Health Organization, there are currently only 26 infectious diseases which are preventable by available vaccines (World Health Organization, 2020).

By developing better risk measures for therapeutic development, we hope to facilitate greater investment, identification of underserved areas that require public-sector support, and more efficient business and financing models in this critical field.

### Methods

We apply the method of Wong, Siah & Lo (2019b) to estimate the PoS of drug development programs using historical clinical trial data. This method was also applied in Wong, Siah & Lo (2019a) to investigate the clinical success rates of oncology development programs. We briefly describe this method, with parts reproduced from the aforementioned papers for expositional convenience.

A drug development program is the clinical investigation of the use of a drug for a disease, typically consisting of sequential clinical trials, separated into phases. We make the assumption that each program must transition from phase 1 to phase 2 to phase 3 to approval. We say that a drug development program has reached phase *i* if it is observed, or can be inferred, that there is at least one trial in phase *i*. The probability of a drug development program transitioning from phase *i* to phase *j* (PoS_*ij*_) can be computed using the simple ratio N_*j*_/N_*i*_, where N_*i*_ is the number of drug development programs initiated at phase *i* (where *i* = 1, 2, or 3) with known outcomes between phase *i* and phase *j* (where *j* = 2, 3, or “A” which denotes regulatory approval), and N_*j*_ is the number of drug development programs among the former that made it to phase *j*.

We call the estimated probability of a drug development program transitioning from phase *i* to phase *i*+1 the “phase *i* PoS”, and the “estimated overall PoS” is defined as the estimated probability of a drug development program going from phase 1 to regulatory approval in at least one country. The estimated probability of a drug development program transitioning from phase 1 to approval—estimated directly using the method described above—is called the “path-by-path” estimate of the overall PoS, and is reported for all PoS calculations.

It should be emphasized that because of this treatment of in-progress drug development programs, path-by-path PoS estimates are not multiplicative, i.e., PoS_12_ x PoS_23_ x PoS_3A_ :;t PoS_1A_, in contrast to phase-by-phase estimates, which do multiply (see Wong, Siah & Lo (2019b) and https://projectalpha.mit.edu/faq for details and illustrative examples).

To simplify this terminology, we will henceforth omit the qualifier “estimated” when referring to the PoS, so it should be understood that all PoS values reported in this article are statistical estimates of unobservable population parameters.

When identifying phase transitions, we make the standard assumption that phase 1/2 and phase 2/3 trials are to be considered as phase 2 and phase 3, respectively. We report only drug development programs that have seen at least one trial with a definite outcome.

### Data

We extracted clinical trials metadata from the January 7, 2020, snapshots of Citeline’s PharmaProjects and TrialTrove databases, provided by Informa Pharma Intelligence. Clinical trial metadata was retrieved from the TrialTrove database while the approval data was obtained from the PharmaProjects database, both of which are required to identify the drug development programs. These databases contain information from both US and non-US sources. We consider a drug approved if it is approved in any country. All clinical trials used in this analysis have end dates after January 1, 2000, and start dates before January 7, 2020.

We filter our data to include only trials that have been tagged by Citeline as being in the ‘Infectious Disease’ or ‘Vaccines (Infectious Diseases)’ therapeutic areas. The vaccine types and diseases are provided by the databases. The database encodes each unique triplet of trial identification number, drug, and disease as a data point. As such, a single trial can be repeated as multiple data points. Since the two therapeutic areas may overlap in data points, we define clinical trials that are involved in any vaccine development as part of a ‘vaccine’ development program. In addition, we process the data such that more specific diseases (e.g., rabies) can be identified instead of broad vaccine classes (e.g., vector-borne disease vaccines). Clinical trials that are not involved in any vaccine development program will be deemed to be part of a ‘non-vaccine’ drug development program. We derive 43,414 data points in total. We define an ‘industry-sponsored’ development program as one where there is at least one commercial company involved in any stage of clinical development. The complement—in which there is no commercial company involved in any stage of the vaccine or drug development program—shall be referred to as ‘non-industry-sponsored’.

We plot the number of development programs known to start in each month from January 2000 through December 2019 in Figure 1. There are 1,838 and 706 industry-sponsored and non-industry-sponsored vaccine development programs, respectively, and, 3,851 and 2,978 industry-sponsored and non-industry-sponsored non-vaccine drug development programs targeting infectious diseases, respectively. As can be seen from Figure 1a, the number of industry-sponsored clinical programs attempting to treat infectious diseases is often greater than the number of vaccine development programs. The gap between the number of programs initiated for each development type seems to have increased between January 2000 and May 2014 before declining (see Section A7 for a more detailed analysis). We see a precipitous fall in the number of infectious disease treatment development programs initiated between late 2018 and mid-2019, which is likely to be related to declining investment in the research and development (R&D) of novel antibiotics, precipitated by the closure of antibiotics biotechnology firms and the withdrawal of pharmaceutical companies from the antibiotics business (Hu, 2018; Langreth, 2019).

**Figure 1.**
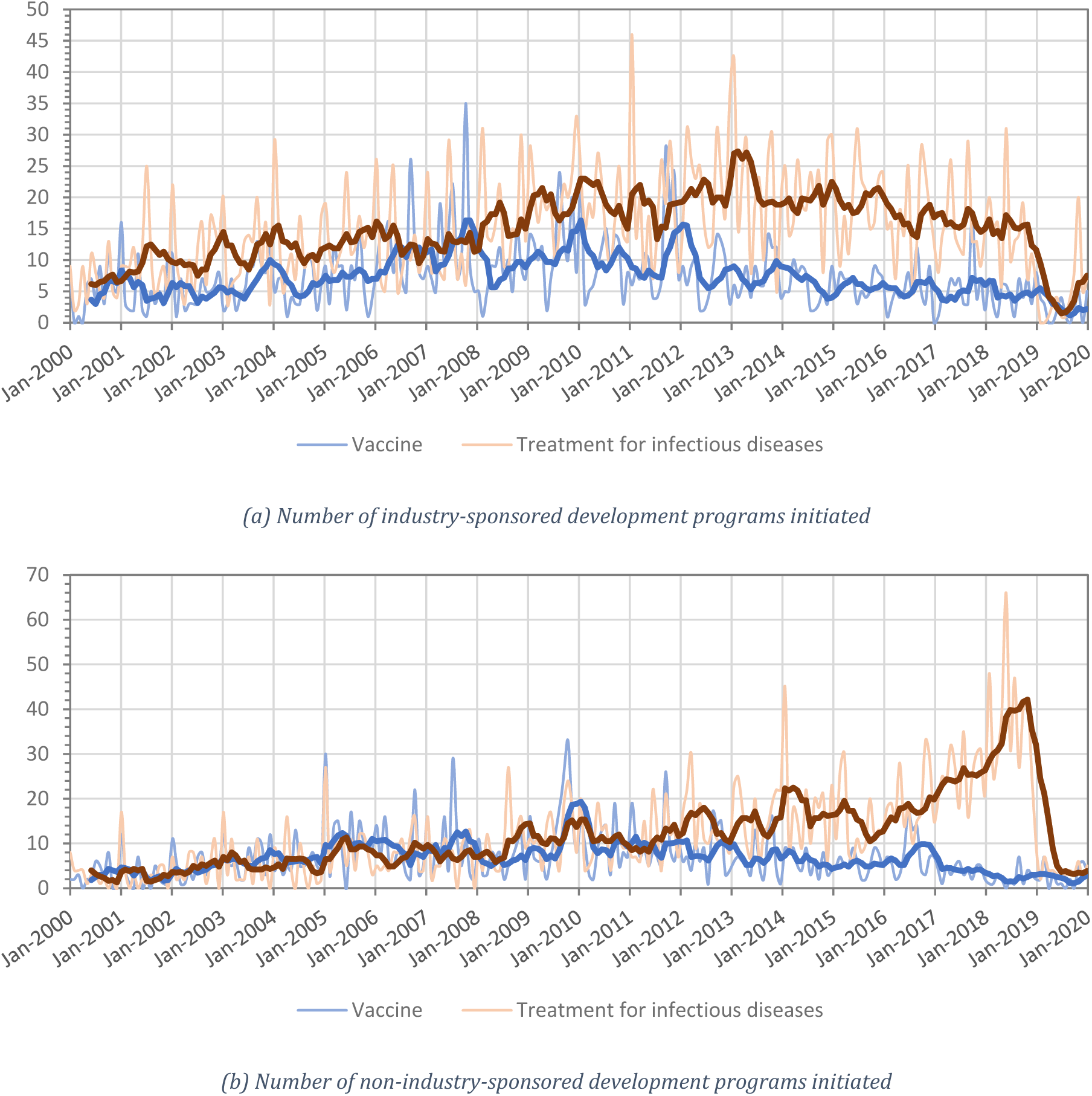
The number of development programs initiated per month from January 2000 through December 2019 in the areas of vaccine and non-vaccine treatment for infectious diseases. The darker, thicker lines represent the 6-month moving average of the individual series.

Between January 2000 and June 2011, the number of non-industry-sponsored vaccine development programs initiated is on par with the number of non-industry-sponsored, non-vaccine anti-infective drug development programs initiated (see Figure 1b). However, the number of non-vaccine drug development programs initiated begin to outpace the number of vaccine development programs after January 2012, and experience a rapid boom between mid-2015 and mid-2018 before declining rapidly between October 2018 and January 2019.

### Results

## Vaccines

Overall, 2,544 vaccine development programs are observed in our dataset, of which 1,838 are sponsored by industry and 706 do not involve any industry sponsor in any stage of development. For industry-sponsored drug development programs, ‘respiratory infections’ is the most actively researched vaccine category, comprising 34.8% (n=640) of all vaccine development programs (see Figure 2). Hepatitis B virus (HBV) and human immunodeficiency virus (HIV) vaccines represent 11.6% (n=213) and 9.8% (n=181) of all vaccine development programs, respectively, whereas intra-abdominal infections, monkey pox, and severe acute respiratory syndrome (SARS) vaccines are the least researched fields, with only one development path observed per disease.

**Figure 2.**
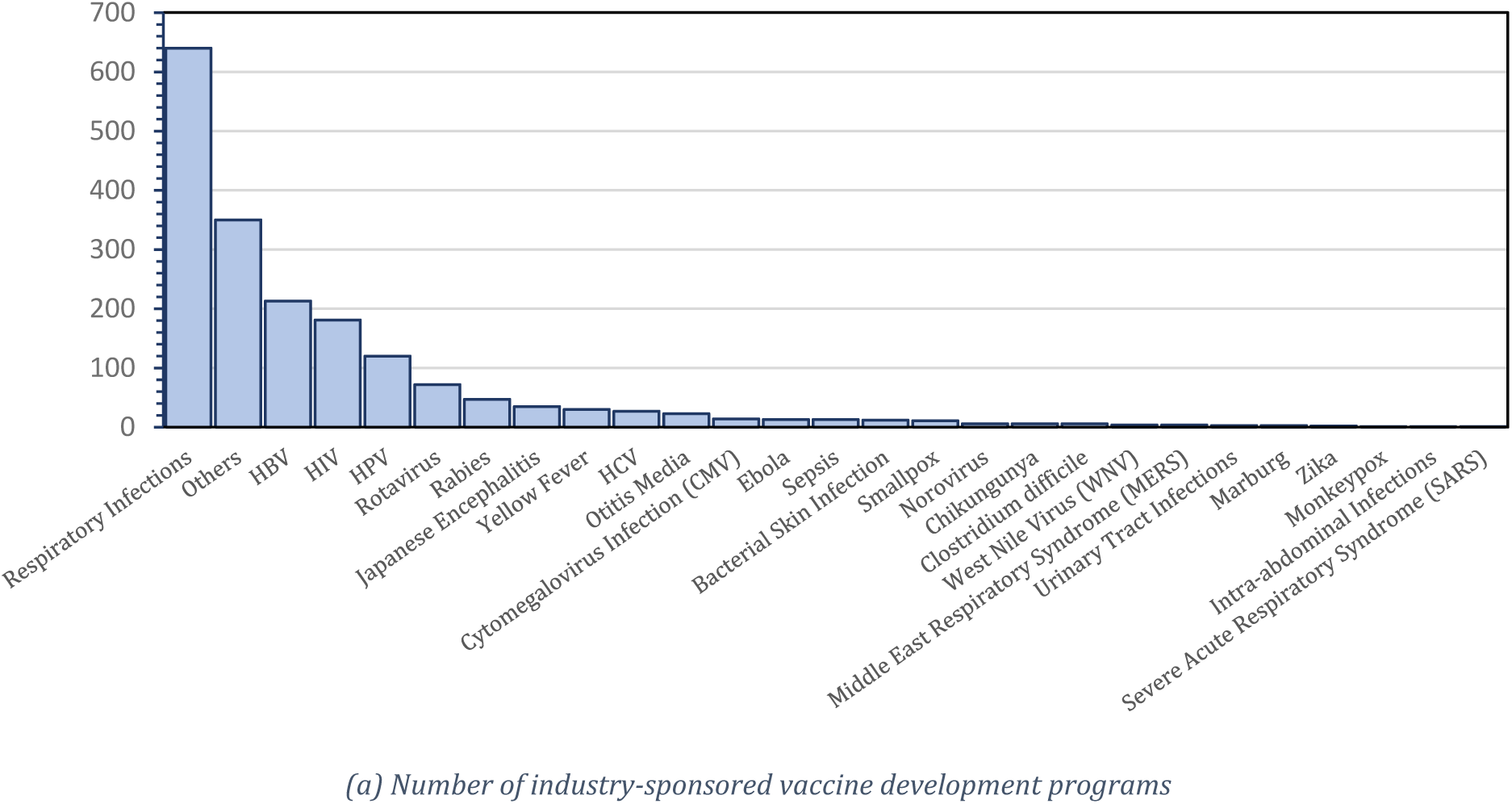
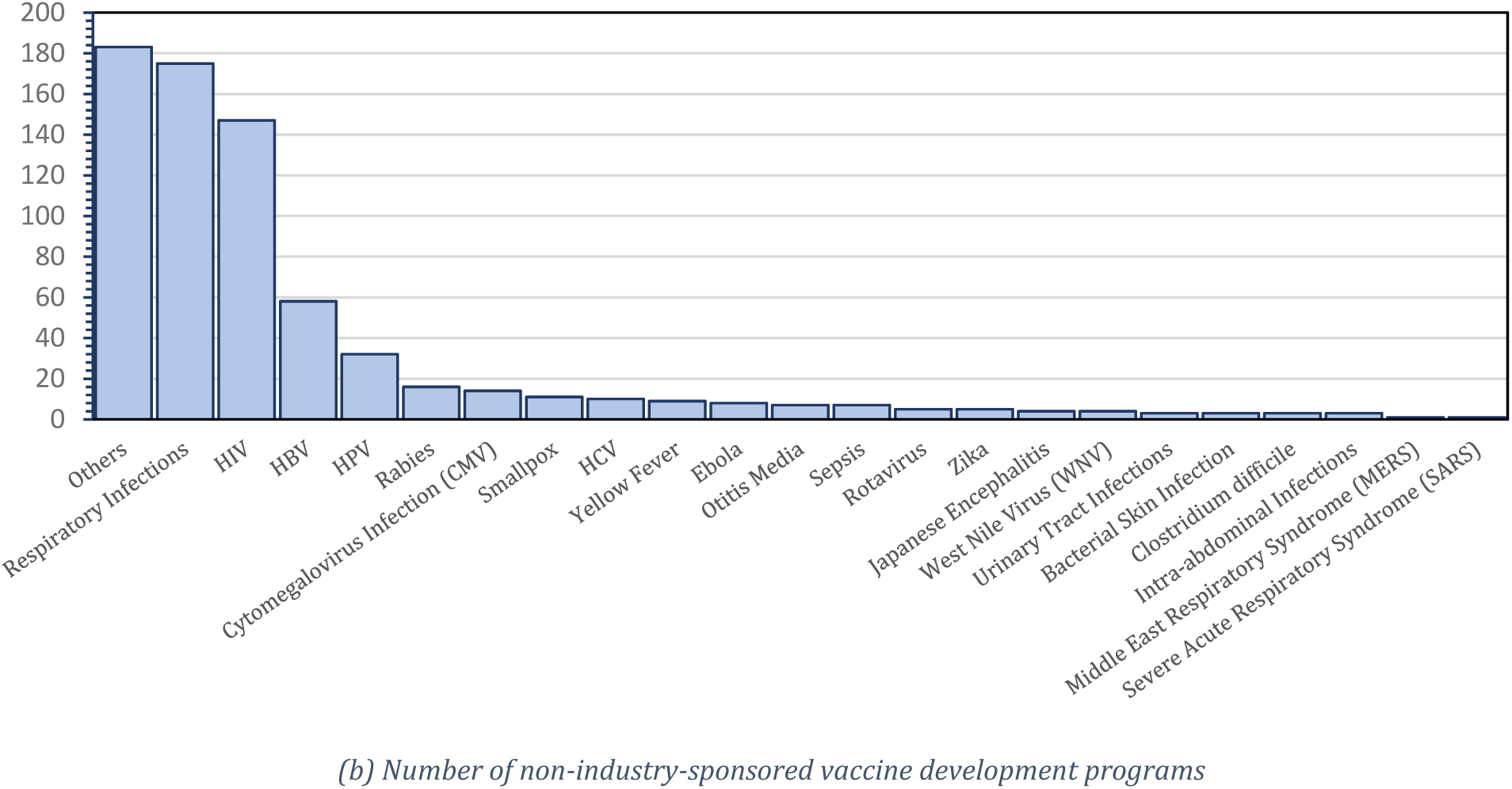
Number of vaccine development programs observed for each vaccine type. HBV, hepatitis B virus; HCV, hepatitis C virus; HIV, human immunodeficiency virus; HPV, human papillomavirus.

A similar pattern can be seen for the non-industry-sponsored vaccine development programs; excluding the ‘others’ category, the top three most researched vaccine categories are also respiratory infections (24.8%), HIV (20.4%), and HBV (8.2%), whereas Middle East Respiratory Syndrome (MERS) and SARS are the least researched diseases with one development program each.

From Figure 3a, we can see that the overall PoS for industry-sponsored vaccine development programs is 39.6% (standard error, or SE: 1.2%), which is substantially higher than the average overall PoS of 11.0% (SE: 0.2%) across all industry-sponsored drug development programs (see Table 2 in the Supplementary Materials). These findings are largely in line with the results of Wong et al. (2019b), who first observed this trend, and of DiMasi et al. (2020), despite the fact that the latter computed their estimates using a different method (a “phase-by-phase” approach) and considered only lead indications. We estimate PoS_12_, PoS_23_, and PoS_3A_ to be 82.5% (SE: 0.9%), 65.4% (SE: 1.3%), and 80.1% (SE: 1.4%), respectively.

**Figure 3.**
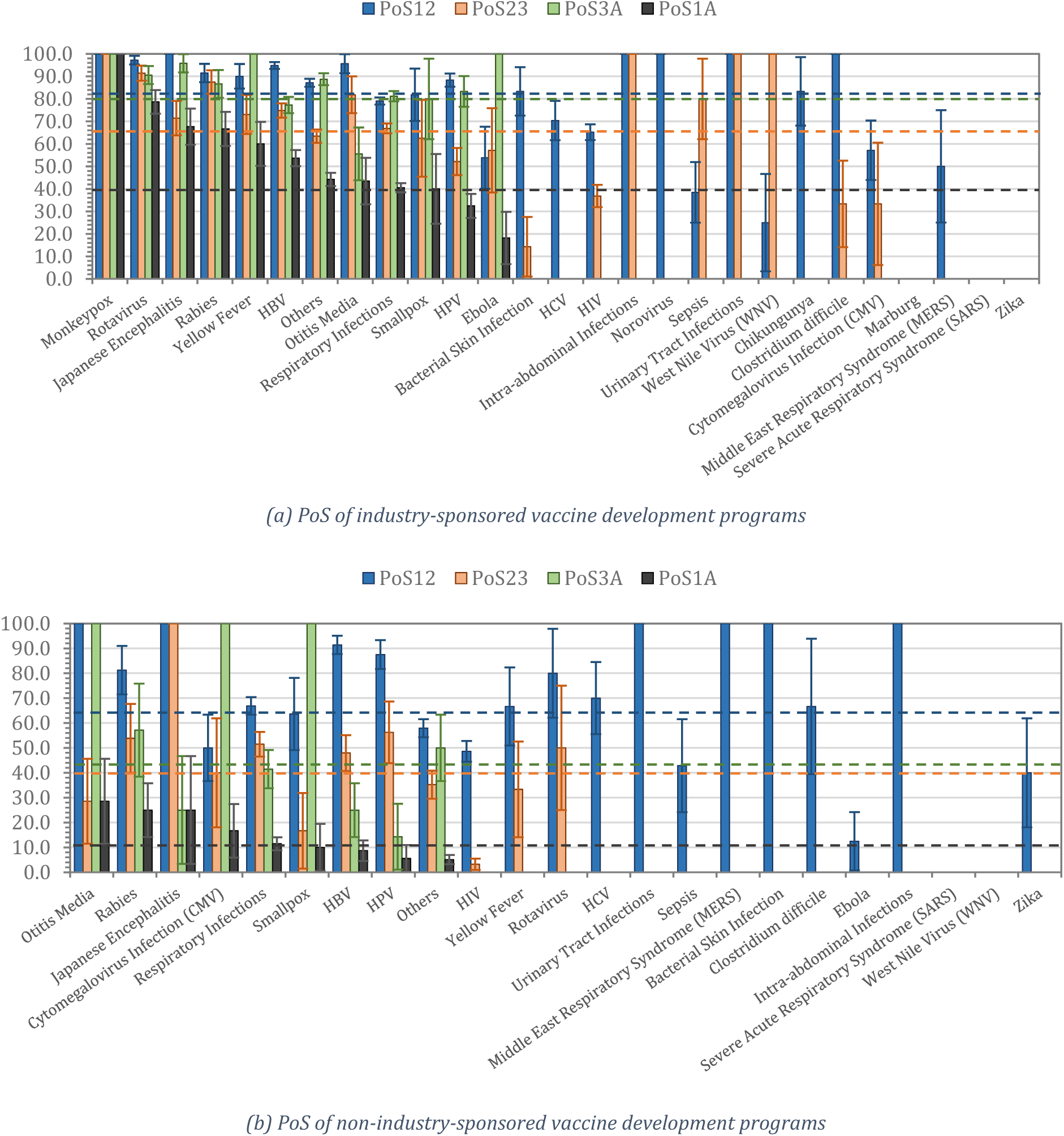
The probabilities of success (PoS) for the different vaccines. The dashed lines represent the overall PoS across all vaccines. PoS_12_, PoS_23_, PoS_3A_, and PoS_1A_ refer to the probability of transitioning from phase 1 to phase 2, phase 2 to phase 3, phase 3 to approval, and phase 1 to approval, respectively. HBV, hepatitis B virus; HCV, hepatitis C virus; HIV, human immunodeficiency virus; HPV, human papillomavirus.

Across all industry-sponsored vaccine development programs, we can see that monkeypox vaccines have had the most developmental success, followed by rotavirus and Japanese encephalitis vaccines (see Figure 3). Their overall success rates are 100% (SE: 0%), 78.7% (SE: 5.2%), and 67.6% (SE: 8.0%), respectively. The PoS for monkeypox is based on only one sample. Only 12 diseases out of the 27 disease categories with at least one development path observed have seen at least one approved vaccine.

In contrast, non-industry-sponsored vaccine development programs have an overall PoS of only 6.8% (SE: 1.0%), with PoS_12_, PoS_23_, and PoS_3A_ estimates of 63.3% (SE: 1.8%), 37.3% (SE: 2.6%), and 39.8% (SE: 4.9%), respectively (Figure 3b). The top three indications with the highest overall success rates for non-industry-sponsored drug development programs are otitis media (28.6%, SE: 17.1%), rabies (25.0%, SE: 10.8%), and Japanese encephalitis (25.0%, SE: 21.7%). The latter estimates are derived from only a handful of samples and must be interpreted with caution as their large standard errors suggest.

### Non-Vaccine Anti-Infective Therapeutics

In contrast to vaccines, which are intended to prevent disease, a number of alternatives have been developed to treat—and, in some cases, cure—patients afflicted with an infectious disease. According to our dataset, 3,851 and 2,978 industry-sponsored and non-industry-sponsored non-vaccine drug development programs, respectively, have been initiated in the area of infectious disease (see Figure 4). The top three diseases with the greatest number of industry-sponsored drug development programs are respiratory infections (21.8%), HIV (15.7%) and hepatitis C virus, or HCV (14.1%). Together, they comprise 51.6% of all industry-sponsored non-vaccine development programs. Non-industry anti-infectious-disease drug development programs focus on treating respiratory infections (20.5%), HIV (13.9%), and bacteria skin infection (12.1%).

**Figure 4.**
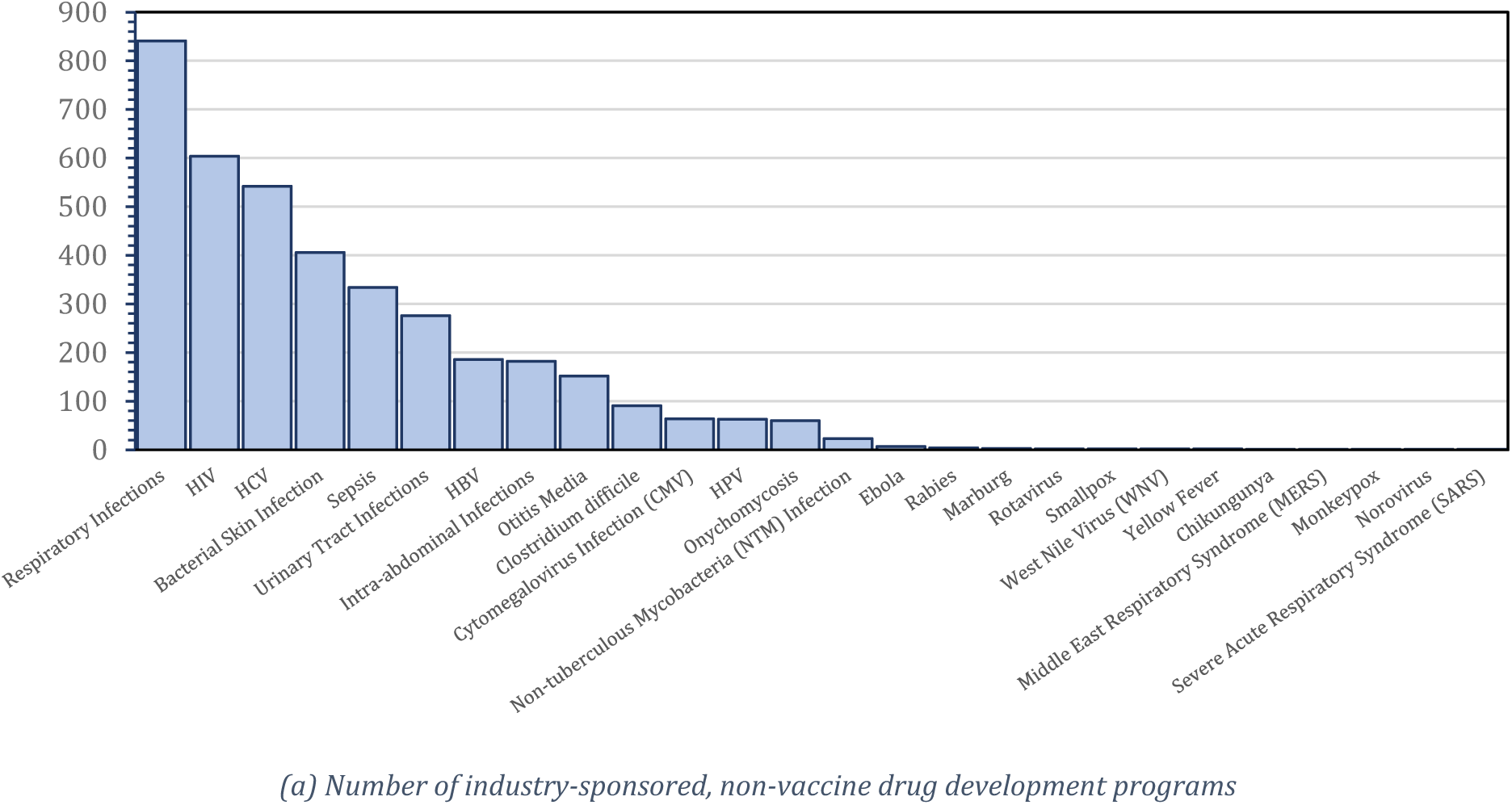
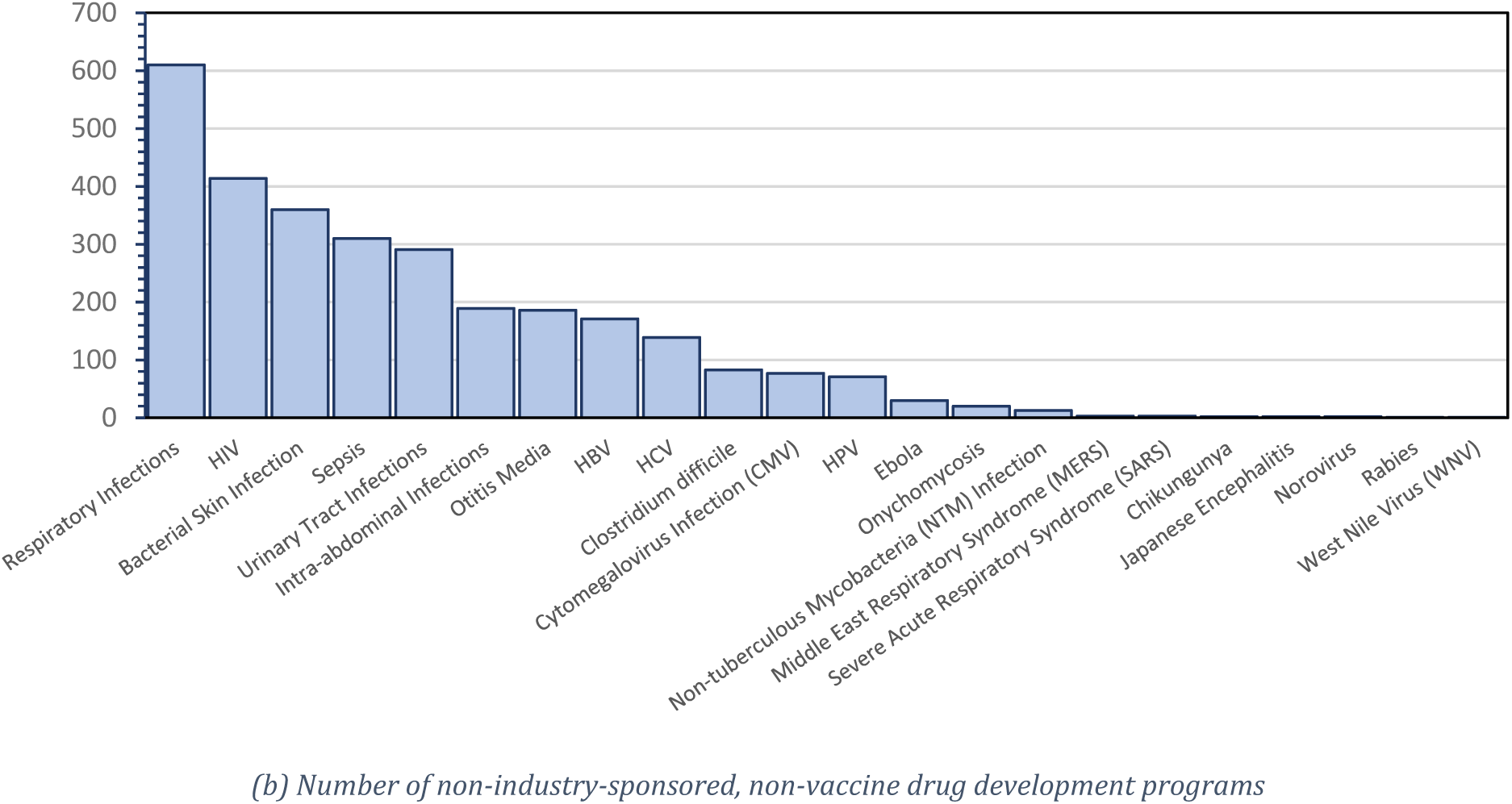
Number of non-vaccine drug development programs for each disease in the ‘Infectious Disease’ category. HBV, hepatitis B virus; HCV, hepatitis C virus; HIV, human immunodeficiency virus; HPV, human papillomavirus.

With respect to addressing the most recent virus outbreaks—MERS, SARS, Ebola, and Zika—a total of 9 industry-sponsored and 36 non-industry-sponsored non-vaccine drug development programs were initiated over the past twenty years, and there have been no approved therapies to date.

From Figure 5a, we can see that the overall PoS across all industry-sponsored drug development programs treating infectious diseases is 16.3% (SE: 0.7%). The PoS_12_, PoS_23_, and PoS_3A_ are 65.0% (SE: 0.8%), 64.3% (SE: 1.0%), and 51.1% (SE: 1.6%), respectively. Based on our data, the highest success rates for industry-sponsored non-vaccine development programs have been for smallpox (100.0%, SE: 0.0%), CMV infection (31.8%, SE: 7.0%), and onychomycosis (29.8%, SE: 6.7%). There are currently no non-vaccine therapies approved for rotavirus, SARS, rabies, Ebola, West Nile Virus (WNV), Marburg, yellow fever, chikungunya, MERS, monkeypox, or norovirus. With the exception of norovirus and MERS, these diseases without any vaccine are predominantly prevalent in non-industrialized nations, and thus represent neglected diseases. It is also interesting that for the latter eight diseases, even the PoS_12_ is low. Since phase 1 trials in the development of anti-infective therapies focus primarily on safety, understanding the pharmacokinetics of the compound, and maximum tolerable dose levels, it can be inferred that the drugs tested are either of high toxicity or lack the necessary characteristics required for optimal absorption, distribution, metabolism, and excretion (ODME), or perhaps failed to advance due to financial constraints.

**Figure 5.**
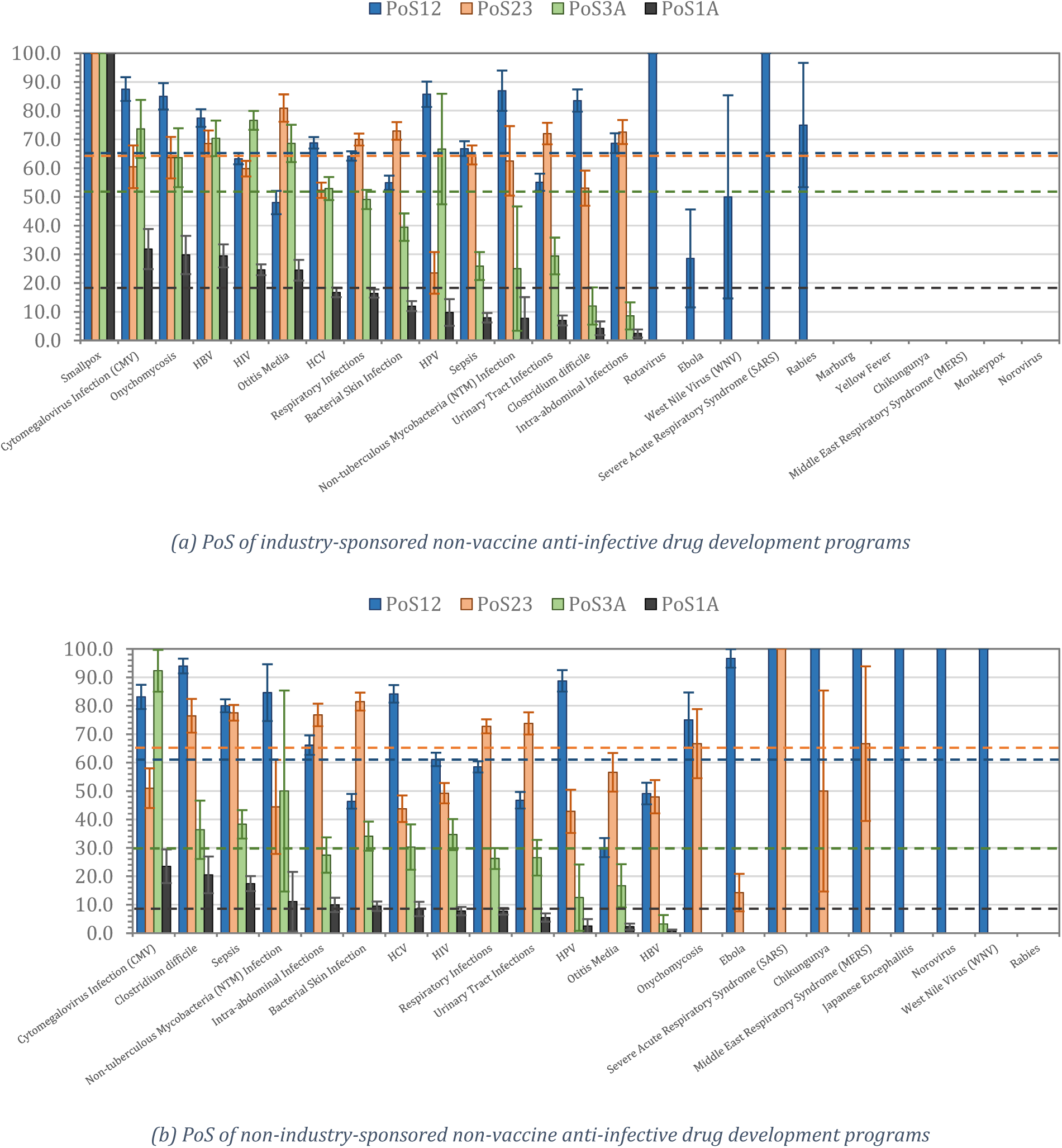
The probabilities of success (PoS) of non-vaccine anti-infective drug development programs for infectious diseases. The dashed lines represent the overall PoS across all diseases. PoS_12_, PoS_23_, PoS_3A_, and PoS_1A_ refer to the probability of transitioning from phase 1 to phase 2, phase 2 to phase 3, phase 3 to approval, and phase 1 to approval, respectively. HBV, hepatitis B virus; HCV, hepatitis C virus; HIV, human immunodeficiency virus; HPV, human papillomavirus.

For non-industry-sponsored non-vaccine development programs, the overall PoS is 8.2% (SE: 0.6%) while PoS_12_, PoS_23_, and PoS_3A_ are 61.0% (SE: 0.9%), 65.2% (SE: 1.2%), and % (SE: 1.8%), respectively (see Figure *5*b). The top three indications with the highest overall success rates for non-industry-sponsored non-vaccine development programs are CMV infection (23.5%, SE: 5.9%), clostridium difficile (20.5%, SE: 6.5%), and sepsis (17.4%, SE: 2.6%).

## Industry-Sponsored Trials

In an attempt to shed more light on the industry-sponsored vaccine and non-vaccine drug development programs, we classify the diseases by their biological family and transmission type. The classifications are presented in Table 1 in the Supplementary Materials. We then compute the PoS using these classifications.

**Table 1.** List of transmission routes and biological family for the infectious diseases

| Disease | Transmission Route | Biological Family |
| --- | --- | --- |
| Hepatitis B virus (HBV) | Human-human | Hepadnavirus |
| Other | Multiple or others | Multiple or others |
| Otitis Media | Multiple or others | Multiple or others |
| Bacterial Skin Infection | Multiple or others | Multiple or others |
| Sepsis | Multiple or others | Multiple or others |
| Human papillomavirus (HPV) | Human-human | Flaviviridae |
| Human immunodeficiency virus (HIV) | Human-human | Retroviridae |
| Intra-abdominal Infections | Multiple or others | Multiple or others |
| Onychomycosis | Multiple or others | Multiple or others |
| Clostridium difficile | Multiple or others | Clostridiaceae |
| Cytomegalovirus (CMV) Infection | Human-human | Herpesviridae |
| Hepatitis C virus (HCV) | Human-human | Flaviviridae |
| Respiratory Infections | Multiple or others | Multiple or others |
| Urinary Tract Infections | Multiple or others | Multiple or others |
| Rotavirus | Human-human | Reoviridae |
| Ebola | Human-human | Filoviridae |
| Marburg | Human-human | Filoviridae |
| Smallpox | Human-human (Airborne) | Poxvirus |
| Zika | Animal bites | Flaviviridae |
| Rabies | Animal bites | Rhabdoviridae |
| Yellow Fever | Animal bites | Flaviviridae |
| Chikungunya | Animal bites | Togaviridae |
| Norovirus | Contaminated food or water | Caliciviridae |
| Japanese Encephalitis | Animal bites | Flaviviridae |
| Non-tuberculous Mycobacteria (NTM) Infection | Multiple or others | Multiple or others |
| West Nile Virus (WNV) | Animal bites | Flaviviridae |
| Middle East Respiratory Syndrome (MERS) | Human-human (Airborne) | Coronaviridae |
| Severe Acute Respiratory Syndrome (SARS) | Human-human (Airborne) | Coronaviridae |
| Monkeypox | Animal bites | Poxviridae |

**Table 2.** The probabilities of success (PoS) of industry-sponsored drug development programs across all therapeutic groups. The classification of vaccines used in this table is based on broader categories such as ‘other viral vaccines’ instead of the finer ones such as ‘ebola’ used in this paper, resulting in a slight difference in the computed PoSs. A: regulatory approval; P1: phase 1; P2: phase 2; P3: phase 3; SE: standard error.

| Disease | P1 Paths | PoS<sub>12</sub> | S.E. | P2 Paths | PoS<sub>23</sub> | S.E. | PoS<sub>2A</sub> | S.E. | P3 Paths | PoS<sub>3A</sub> | S.E. | PoS<sub>1A</sub> | S.E. |
| --- | --- | --- | --- | --- | --- | --- | --- | --- | --- | --- | --- | --- | --- |
| Oncology | 27,600 | 65.5 | 0.3 | 10,650 | 37.7 | 0.5 | 6.9 | 0.3 | 2,597 | 28.2 | 0.9 | 3.9 | 0.1 |
| Metabolic/Endocrinology | 4,360 | 74.2 | 0.7 | 2,767 | 60.0 | 0.9 | 21.3 | 0.8 | 1,293 | 45.6 | 1.4 | 16.7 | 0.6 |
| Cardiovascular | 3,387 | 74.3 | 0.8 | 2,265 | 70.2 | 1.0 | 25.9 | 1.0 | 1,203 | 48.8 | 1.4 | 21.4 | 0.8 |
| CNS | 6,207 | 71.7 | 0.6 | 3,806 | 56.9 | 0.8 | 14.6 | 0.6 | 1,525 | 36.5 | 1.2 | 11.3 | 0.5 |
| Autoimmune/Inflammation | 6,272 | 71.7 | 0.6 | 3,704 | 52.7 | 0.8 | 17.3 | 0.7 | 1,332 | 48.0 | 1.4 | 13.1 | 0.5 |
| Genitourinary | 1,103 | 71.4 | 1.4 | 737 | 61.6 | 1.8 | 25.0 | 1.7 | 352 | 52.3 | 2.7 | 19.3 | 1.3 |
| Infectious Disease (ex. vaccines) | 3,851 | 65.0 | 0.8 | 2,202 | 64.2 | 1.0 | 23.1 | 1.0 | 996 | 51.0 | 1.6 | 16.2 | 0.7 |
| Ophthalmology | 697 | 89.1 | 1.2 | 510 | 55.7 | 2.2 | 17.1 | 1.8 | 191 | 45.5 | 3.6 | 17.6 | 1.7 |
| Vaccines (Infectious Disease) | 1,886 | 83.9 | 0.8 | 1,409 | 66.4 | 1.3 | 45.8 | 1.4 | 813 | 79.5 | 1.4 | 40.6 | 1.2 |
| Total | 55,363 | 69.1 | 0.2 | 28,050 | 51.6 | 0.3 | 16.1 | 0.2 | 10,302 | 44.0 | 0.5 | 11.0 | 0.2 |
| All except oncology | 27,763 | 72.7 | 0.3 | 17,400 | 60.1 | 0.4 | 21.8 | 0.3 | 7,705 | 49.3 | 0.6 | 17.1 | 0.3 |

Looking at the vaccine PoS by transmission route (see Figure 6a), we see that vaccines for diseases transmitted through animal bites have the highest overall PoS (61.3%, SE: 4.7%), whereas no vaccine has been developed for diseases transmitted through contaminated food or water.

**Figure 6.**
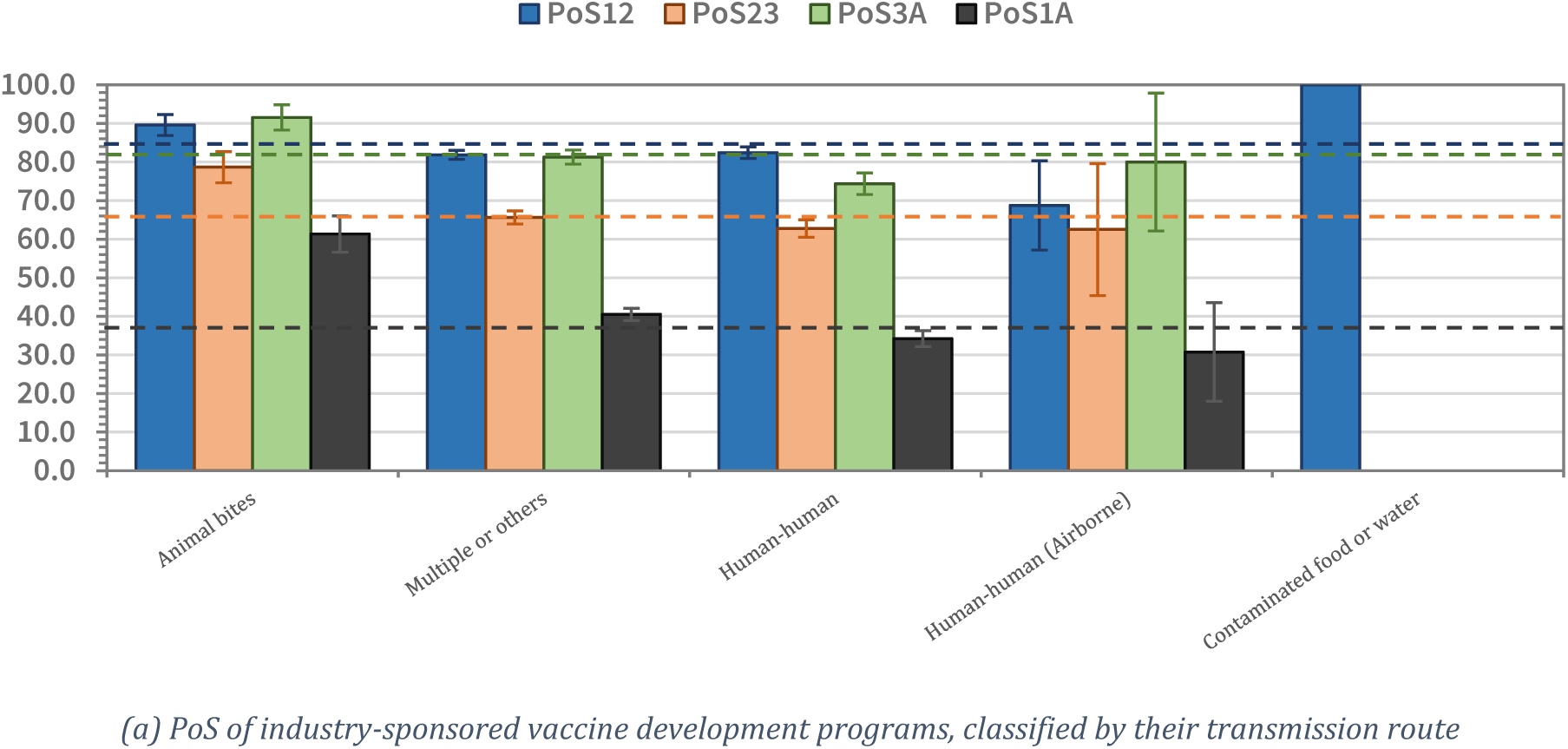
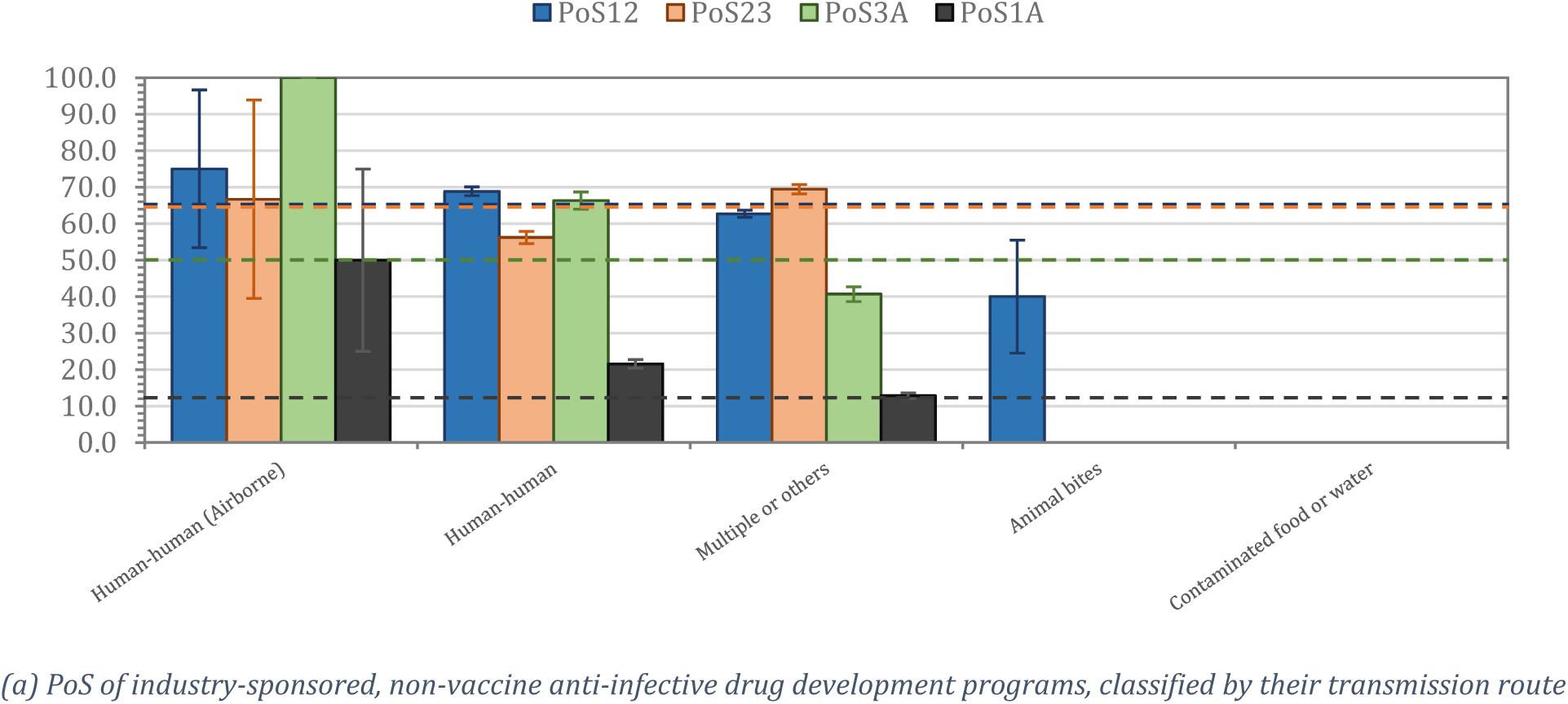
The probabilities of success (PoS) of industry-sponsored vaccine and non-vaccine anti-infective drug development programs, classified by their transmission route. The dashed lines represent the overall PoS in the disease transmission type. PoS_12_, PoS_23_, PoS_3A_, and PoS_1A_ refer to the probability of transitioning from phase 1 to phase 2, phase 2 to phase 3, phase 3 to approval, and phase 1 to approval, respectively.

We find that companies have been most successful in developing non-vaccine treatments for diseases transmitted between humans through the air, with 50.0% (SE: 25.0%) of all drug development programs making it from phase 1 to regulatory approval (see Figure 6b). Unfortunately, this is based on only four drug development programs and may not be indicative of the general trend. Treatments for diseases that transmit through ‘human to human (others)’ have an overall PoS of 21.5% (SE: 1.2%) while no approval is observed for diseases transmitted through ‘animal bites’ or ‘contaminated food or water’.

When we classify the vaccines by the biological family of the infectious agent (Figure 7a), we see that *reoviridae* (e.g., rotavirus), *rhabdoviridae* (e.g., rabies), and *hepadnaviridae* (e.g., HBV) are the three biological families with the highest overall PoS for vaccines at 78.7%, (SE: 5.2%), 66.7% (SE: 7.5%), and 53.7% (SE: 3.6%), respectively. We have yet to see a vaccine for diseases caused by agents in the biological families of *retroviridae* (e.g., HIV), *caliciviridae* (e.g., norovirus), *clostridiaceae* (e.g., clostridium difficile), *coronaviridae* (e.g., SARS, MERS), *herpesviridae* (e.g., CMV infection), or *togaviridae* (e.g., chikungunya).

**Figure 7.**
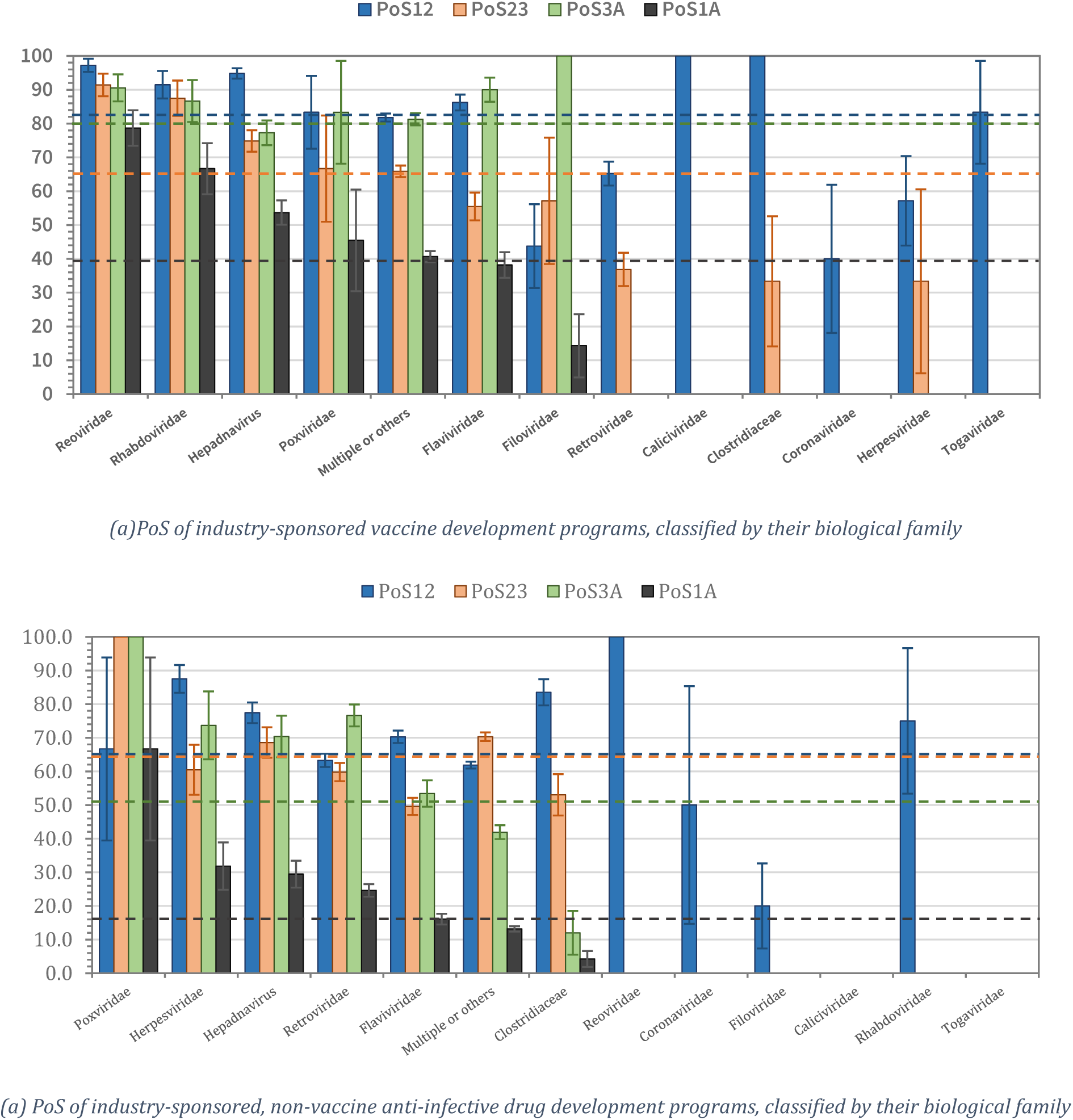
The probabilities of success (PoS) of vaccine and non-vaccine anti-infective drug development programs, classified by their biological family. The dashed lines represent the overall PoS across all diseases. PoS_12_, PoS_23_, PoS_3A_, and PoS_1A_ refer to the probability of transitioning from phase 1 to phase 2, phase 2 to phase 3, phase 3 to approval, and phase 1 to approval, respectively.

**Figure 8.**
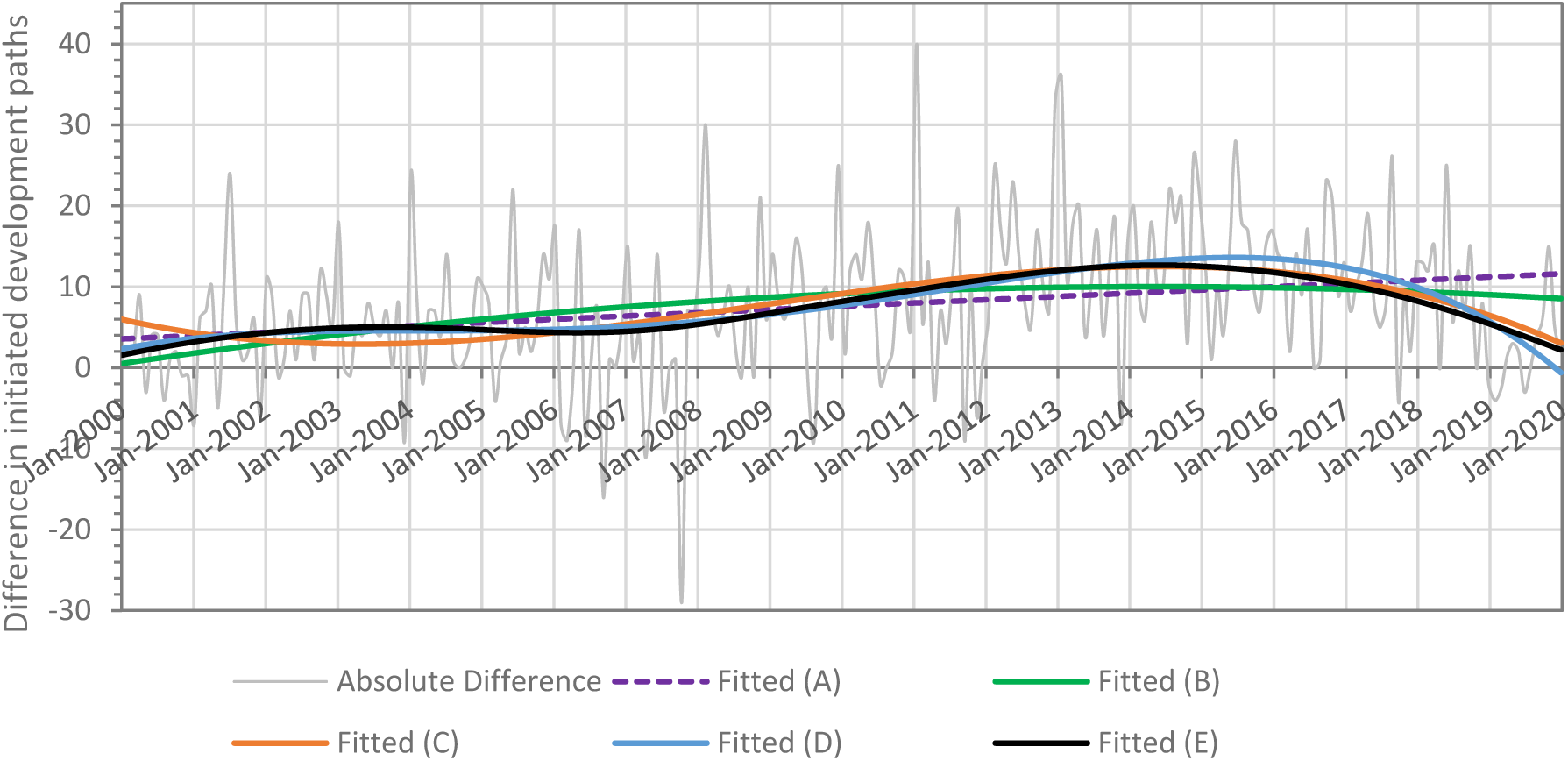
Time series of the difference between the number of investigational non-vaccine treatments initiated and investigational vaccines initiated, and the fitted lines for the regression models.

When we consider non-vaccine PoS by biological family of the infectious agent (see Figure 7b), we see that non-vaccine therapies for *poxviridae* (e.g., smallpox), *herpesviridae* (e.g., CMV infection), and *hepadnaviridae* (e.g., HBV) have the highest overall PoS at 66.7% (SE: 27.2%), 31.8% (SE: 7.0%), and 29.5% (SE: 4.0%), respectively. For viruses in the *reoviridae* (e.g., rotavirus), *coronaviridae* (e.g., SARS, MERS), *caliciviridae* (e.g., norovirus), *rhabdoviridae* (e.g., rabies), and *togaviridae* (e.g., chikungunya) families, there have been less than five development programs each, and no approved treatments.

## Discussion

Companies producing vaccines and other therapeutics for infectious diseases have gradually been retreating from these spaces in recent years. The number of companies producing vaccines has dwindled over the past few decades, and the top four vaccine companies now make up more than 90% of the global market (Evaluate, 2018). Similarly, the top four companies producing antiviral drugs occupy about 80% of the global market (Evaluate, 2018). Antibiotic developers such Achaogen and Melinta Therapeutics have filed for bankruptcy in the past year, while large pharmaceutical companies such as Novartis and Sanofi have withdrawn from the space (Jacobs, 2019), leading the Infectious Diseases Society of America to sound the alarm about the availability of effective antibiotics (Infectious Diseases Society of America, 2019).

It should be no surprise that investors are unwilling to invest in companies producing vaccines and treatments for infectious diseases given the economics of this market (Vu et al., 2020). These have been generally regarded as low-margin products, and they have low growth potential compared to treatments in other therapeutic areas, such as oncology or cardiovascular diseases. As a comparison, Revlimid, the blockbuster cancer drug for multiple myeloma, earned $9.69 billion for Celgene in 2019 (Celgene, 2019), whereas the vaccine and antiviral portfolios of GlaxoSmithKline, the top vaccine producer by sales, generated only $6.65 billion and $5.89 billion in revenues, respectively, in 2017 (Evaluate, 2018).

This lack of investment has resulted in a relatively low number of development programs for vaccines and treatments of infectious diseases. Only 5,689 industry-sponsored programs were initiated in the past two decades, a mere 12.5% of all industry-sponsored drug development programs launched in the same period.

Our study indicates that the technical success rate is unlikely to be a barrier to investments in new vaccines and treatments for infectious diseases, unlike cancer drugs, where the financial risk of new R&D projects comes from the reduced chance of bringing a drug-indication pair from phase 1 to market. The overall PoS of industry-sponsored vaccines and treatments for infectious diseases are above the average for all therapeutic groups (see Table 2 in the Supplementary Materials).

It is often suggested that the fundamental issue behind this lack of investment is that the market for vaccines and treatments for infectious diseases is simply not lucrative enough. Despite the expense of research and development and the need for large-scale production (Weir & Gruber, 2016), anti-infective disease treatments are used only occasionally, while vaccine companies face an avalanche of liability lawsuits (Hensley & Wysocki Jr., 2005). Furthermore, the companies are at the mercy of government pricing decisions (Hu, 2018).

It remains to be seen if more non-industry-sponsored research can alleviate the issue. Our study shows that only 6.8% (SE: 1.0%) and 8.2% (SE: 0.6%) of non-industry-sponsored vaccines and non-vaccine infectious disease development programs transition from phase 1 to approval, respectively. However, this may be a result of selection bias: promising vaccine and therapeutics initiated in non-industry settings are often pursued in conjunction with industry-sponsored sponsors, whereas commercially less promising projects are more likely to be pursued by non-profit organizations.

## Conclusion

The world today has never been in greater need of effective vaccines and other anti-infectives. As the COVID-19 crisis has shown, infectious diseases still have the potential to cause a catastrophically large number of deaths and disrupt the daily lives of billions. We hope that our research into the probability of successfully developing infectious disease therapeutics will inform all the stakeholders and catalyze innovation and greater investment in this critical and underserved field.

## Data Availability

All data used are commercially available datasets that other authors can access on the same terms.

https://informa.com

## Supplementary Materials

### A1. List of transmission route and biological family for infectious diseases

### A2. PoS Tables across all therapeutic areas

We reproduce the probability of success across all therapeutic groups.

### A3. PoS Tables for vaccines (industry-sponsored)

**Table 3.** The probabilities of success (PoS) of industry-sponsored vaccine drug development programs. A: regulatory approval; P1: phase 1; P2: phase 2; P3: phase 3; SE: standard error.

| Disease | P1 Paths | PoS<sub>12</sub> | S.E. | P2 Paths | PoS<sub>23</sub> | S.E. | PoS<sub>2A</sub> | S.E. | P3 Paths | PoS<sub>3A</sub> | S.E. | PoS<sub>1A</sub> | S.E. |
| --- | --- | --- | --- | --- | --- | --- | --- | --- | --- | --- | --- | --- | --- |
| Bacterial Skin Infection | 12 | 83.3 | 10.8 | 7 | 14.3 | 13.2 | 0.0 | 0.0 | 1 | 0.0 | 0.0 | 0.0 | 0.0 |
| Chikungunya | 6 | 83.3 | 15.2 | 0 | - | - | - | - | 0 | - | - | 0.0 | 0.0 |
| Clostridium difficile | 6 | 100.0 | 0.0 | 6 | 33.3 | 19.2 | 0.0 | 0.0 | 0 | - | - | 0.0 | 0.0 |
| Cytomegalovirus Infection (CMV) | 14 | 57.1 | 13.2 | 3 | 33.3 | 27.2 | 0.0 | 0.0 | 0 | - | - | 0.0 | 0.0 |
| Ebola | 13 | 53.8 | 13.8 | 7 | 57.1 | 18.7 | 28.6 | 20.2 | 2 | 100.0 | 0.0 | 18.2 | 11.6 |
| Hepatitis B Virus (HBV) | 213 | 94.8 | 1.5 | 187 | 74.9 | 3.2 | 54.5 | 3.7 | 132 | 77.3 | 3.6 | 53.7 | 3.6 |
| Hepatitis C Virus (HCV) | 27 | 70.4 | 8.8 | 15 | 0.0 | 0.0 | 0.0 | 0.0 | 0 | - | - | 0.0 | 0.0 |
| Human Immunodeficiency Virus (HIV) | 181 | 65.2 | 3.5 | 95 | 36.8 | 4.9 | 0.0 | 0.0 | 21 | 0.0 | 0.0 | 0.0 | 0.0 |
| Human Papillomavirus (HPV) | 120 | 88.3 | 2.9 | 69 | 52.2 | 6.0 | 36.2 | 6.1 | 30 | 83.3 | 6.8 | 32.5 | 5.3 |
| Intra-abdominal Infections | 1 | 100.0 | 0.0 | 1 | 100.0 | 0.0 | 0.0 | 0.0 | 1 | 0.0 | 0.0 | 0.0 | 0.0 |
| Japanese Encephalitis | 35 | 100.0 | 0.0 | 35 | 71.4 | 7.6 | 65.7 | 8.1 | 24 | 95.8 | 4.1 | 67.6 | 8.0 |
| Marburg | 3 | 0.0 | 0.0 | 0 | - | - | - | - | 0 | - | - | 0.0 | 0.0 |
| Middle East Respiratory Syndrome (MERS) | 4 | 50.0 | 25.0 | 0 | - | - | - | - | 0 | - | - | 0.0 | 0.0 |
| Monkeypox | 1 | 100.0 | 0.0 | 1 | 100.0 | 0.0 | 100.0 | 0.0 | 1 | 100.0 | 0.0 | 100.0 | 0.0 |
| Norovirus | 6 | 100.0 | 0.0 | 5 | 0.0 | 0.0 | 0.0 | 0.0 | 0 | - | - | 0.0 | 0.0 |
| Otitis Media | 23 | 95.7 | 4.3 | 22 | 81.8 | 8.2 | 45.5 | 10.6 | 18 | 55.6 | 11.7 | 43.5 | 10.3 |
| Rabies | 47 | 91.5 | 4.1 | 40 | 87.5 | 5.2 | 65.0 | 8.1 | 30 | 86.7 | 6.2 | 66.7 | 7.5 |
| Respiratory Infections | 640 | 79.1 | 1.6 | 465 | 66.9 | 2.2 | 50.1 | 2.4 | 287 | 81.2 | 2.3 | 40.5 | 2.0 |
| Rotavirus | 72 | 97.2 | 1.9 | 70 | 91.4 | 3.3 | 68.6 | 6.0 | 53 | 90.6 | 4.0 | 78.7 | 5.2 |
| Sepsis | 13 | 38.5 | 13.5 | 5 | 80.0 | 17.9 | 0.0 | 0.0 | 4 | 0.0 | 0.0 | 0.0 | 0.0 |
| Severe Acute Respiratory Syndrome (SARS) | 1 | 0.0 | 0.0 | 0 | - | - | - | - | 0 | - | - | 0.0 | 0.0 |
| Smallpox | 11 | 81.8 | 11.6 | 8 | 62.5 | 17.1 | 50.0 | 17.7 | 5 | 80.0 | 17.9 | 40.0 | 15.5 |
| Urinary Tract Infections | 3 | 100.0 | 0.0 | 3 | 100.0 | 0.0 | 0.0 | 0.0 | 1 | 0.0 | 0.0 | 0.0 | 0.0 |
| West Nile Virus (WNV) | 4 | 25.0 | 21.7 | 1 | 100.0 | 0.0 | 0.0 | 0.0 | 1 | 0.0 | 0.0 | 0.0 | 0.0 |
| Yellow Fever | 30 | 90.0 | 5.5 | 26 | 73.1 | 8.7 | 57.7 | 10.5 | 15 | 100.0 | 0.0 | 60.0 | 9.8 |
| Zika | 2 | 0.0 | 0.0 | 0 | - | - | - | - | 0 | - | - | 0.0 | 0.0 |
| Others | 350 | 87.1 | 1.8 | 268 | 63.4 | 2.9 | 47.0 | 3.2 | 142 | 88.7 | 2.7 | 44.2 | 2.9 |
| Total | 1,838 | 82.5 | 0.9 | 1,339 | 65.4 | 1.3 | 45.9 | 1.4 | 768 | 80.1 | 1.4 | 39.6 | 1.2 |

**Table 4.** The probabilities of success (PoS) of industry-sponsored vaccine development programs, grouped by the biological family. A: regulatory approval; P1: phase 1; P2: phase 2; P3: phase 3; SE: standard error.

| Disease | P1 Paths | PoS<sub>12</sub> | S.E. | P2 Paths | PoS<sub>23</sub> | S.E. | PoS<sub>2A</sub> | S.E. | P3 Paths | PoS<sub>3A</sub> | S.E. | PoS<sub>1A</sub> | S.E. |
| --- | --- | --- | --- | --- | --- | --- | --- | --- | --- | --- | --- | --- | --- |
| Caliciviridae | 6 | 100.0 | 0.0 | 5 | 0.0 | 0.0 | 0.0 | 0.0 | 0 | - | - | 0.0 | 0.0 |
| Clostridiaceae | 6 | 100.0 | 0.0 | 6 | 33.3 | 19.2 | 0.0 | 0.0 | 0 | - | - | 0.0 | 0.0 |
| Coronaviridae | 5 | 40.0 | 21.9 | 0 | - | - | - | - | 0 | - | - | 0.0 | 0.0 |
| Filoviridae | 16 | 43.8 | 12.4 | 7 | 57.1 | 18.7 | 28.6 | 20.2 | 2 | 100.0 | 0.0 | 14.3 | 9.4 |
| Flaviviridae | 218 | 86.2 | 2.3 | 146 | 55.5 | 4.1 | 43.2 | 4.3 | 70 | 90.0 | 3.6 | 38.2 | 3.8 |
| Hepadnaviridae | 213 | 94.8 | 1.5 | 187 | 74.9 | 3.2 | 54.5 | 3.7 | 132 | 77.3 | 3.6 | 53.7 | 3.6 |
| Herpesviridae | 14 | 57.1 | 13.2 | 3 | 33.3 | 27.2 | 0.0 | 0.0 | 0 | - | - | 0.0 | 0.0 |
| Multiple or others | 1,042 | 81.8 | 1.2 | 771 | 65.9 | 1.7 | 47.9 | 1.9 | 454 | 81.3 | 1.8 | 40.7 | 1.6 |
| Poxviridae | 12 | 83.3 | 10.8 | 9 | 66.7 | 15.7 | 55.6 | 16.6 | 6 | 83.3 | 15.2 | 45.5 | 15.0 |
| Reoviridae | 72 | 97.2 | 1.9 | 70 | 91.4 | 3.3 | 68.6 | 6.0 | 53 | 90.6 | 4.0 | 78.7 | 5.2 |
| Retroviridae | 181 | 65.2 | 3.5 | 95 | 36.8 | 4.9 | 0.0 | 0.0 | 21 | 0.0 | 0.0 | 0.0 | 0.0 |
| Rhabdoviridae | 47 | 91.5 | 4.1 | 40 | 87.5 | 5.2 | 65.0 | 8.1 | 30 | 86.7 | 6.2 | 66.7 | 7.5 |
| Togaviridae | 6 | 83.3 | 15.2 | 0 | - | - | - | - | 0 | - | - | 0.0 | 0.0 |
| Total | 1,838 | 82.5 | 0.9 | 1,339 | 65.4 | 1.3 | 45.9 | 1.4 | 768 | 80.1 | 1.4 | 39.6 | 1.2 |

**Table 5.** The probabilities of success (PoS) of industry-sponsored vaccine development programs, grouped by the transmission route. A: regulatory approval; P1: phase 1; P2: phase 2; P3: phase 3; SE: standard error.

| Disease | P1 Paths | PoS<sub>12</sub> | S.E. | P2 Paths | PoS<sub>23</sub> | S.E. | PoS<sub>2A</sub> | S.E. | P3 Paths | PoS<sub>3A</sub> | S.E. | PoS<sub>1A</sub> | S.E. |
| --- | --- | --- | --- | --- | --- | --- | --- | --- | --- | --- | --- | --- | --- |
| Animal bites | 125 | 89.6 | 2.7 | 103 | 78.6 | 4.0 | 63.1 | 5.0 | 71 | 91.5 | 3.3 | 61.3 | 4.7 |
| Contaminated food or water | 6 | 100.0 | 0.0 | 5 | 0.0 | 0.0 | 0.0 | 0.0 | 0 | - | - | 0.0 | 0.0 |
| Human-human | 643 | 82.4 | 1.5 | 446 | 62.8 | 2.3 | 39.7 | 2.4 | 238 | 74.4 | 2.8 | 34.2 | 2.1 |
| Human-human (Airborne) | 16 | 68.8 | 11.6 | 8 | 62.5 | 17.1 | 50.0 | 17.7 | 5 | 80.0 | 17.9 | 30.8 | 12.8 |
| Multiple or others | 1,048 | 81.9 | 1.2 | 777 | 65.6 | 1.7 | 47.5 | 1.9 | 454 | 81.3 | 1.8 | 40.5 | 1.6 |
| Total | 1,838 | 82.5 | 0.9 | 1,339 | 65.4 | 1.3 | 45.9 | 1.4 | 768 | 80.1 | 1.4 | 39.6 | 1.2 |

### A4. PoS Tables for non-vaccine anti-infective therapeutics (industry-sponsored)

**Table 6.** The probabilities of success (PoS) of industry-sponsored, non-vaccine anti-infective drug development programs for the treatment of infectious diseases. A: regulatory approval; P1: phase 1; P2: phase 2; P3: phase 3; SE: standard error.

| Disease | P1 Paths | PoS<sub>12</sub> | S.E. | P2 Paths | PoS<sub>23</sub> | S.E. | PoS<sub>2A</sub> | S.E. | P3 Paths | PoS<sub>3A</sub> | S.E. | PoS<sub>1A</sub> | S.E. |
| --- | --- | --- | --- | --- | --- | --- | --- | --- | --- | --- | --- | --- | --- |
| Bacterial Skin Infection | 406 | 54.9 | 2.5 | 207 | 72.9 | 3.1 | 19.8 | 3.2 | 104 | 39.4 | 4.8 | 12.0 | 1.8 |
| Chikungunya | 1 | 0.0 | 0.0 | 0 | - | - | - | - | 0 | - | - | 0.0 | 0.0 |
| Clostridium difficile | 91 | 83.5 | 3.9 | 66 | 53.0 | 6.1 | 4.5 | 2.8 | 25 | 12.0 | 6.5 | 4.2 | 2.4 |
| Cytomegalovirus Infection (CMV) | 64 | 87.5 | 4.1 | 43 | 60.5 | 7.5 | 32.6 | 7.8 | 19 | 73.7 | 10.1 | 31.8 | 7.0 |
| Ebola | 7 | 28.6 | 17.1 | 1 | 0.0 | 0.0 | 0.0 | 0.0 | 0 | - | - | 0.0 | 0.0 |
| Hepatitis B Virus (HBV) | 186 | 77.4 | 3.1 | 105 | 68.6 | 4.5 | 36.2 | 5.2 | 54 | 70.4 | 6.2 | 29.5 | 4.0 |
| Hepatitis C Virus (HCV) | 542 | 68.8 | 2.0 | 348 | 52.3 | 2.7 | 23.6 | 2.4 | 155 | 52.9 | 4.0 | 16.7 | 1.7 |
| Human Immunodeficiency Virus (HIV) | 604 | 63.2 | 2.0 | 326 | 59.8 | 2.7 | 39.3 | 2.8 | 167 | 76.6 | 3.3 | 24.6 | 1.9 |
| Human Papillomavirus (HPV) | 63 | 85.7 | 4.4 | 34 | 23.5 | 7.3 | 11.8 | 5.7 | 6 | 66.7 | 19.2 | 9.8 | 4.6 |
| Intra-abdominal Infections | 182 | 68.7 | 3.4 | 113 | 72.6 | 4.2 | 2.7 | 2.0 | 35 | 8.6 | 4.7 | 2.4 | 1.4 |
| Marburg | 3 | 0.0 | 0.0 | 0 | - | - | - | - | 0 | - | - | 0.0 | 0.0 |
| Middle East Respiratory Syndrome (MERS) | 1 | 0.0 | 0.0 | 0 | - | - | - | - | 0 | - | - | 0.0 | 0.0 |
| Monkeypox | 1 | 0.0 | 0.0 | 0 | - | - | - | - | 0 | - | - | 0.0 | 0.0 |
| Non-tuberculous Mycobacteria (NTM) Infection | 23 | 87.0 | 7.0 | 16 | 62.5 | 12.1 | 6.3 | 7.7 | 4 | 25.0 | 21.7 | 7.7 | 7.4 |
| Norovirus | 1 | 0.0 | 0.0 | 0 | - | - | - | - | 0 | - | - | 0.0 | 0.0 |
| Onychomycosis | 60 | 85.0 | 4.6 | 44 | 63.6 | 7.3 | 31.8 | 7.6 | 22 | 63.6 | 10.3 | 29.8 | 6.7 |
| Otitis Media | 152 | 48.0 | 4.1 | 68 | 80.9 | 4.8 | 51.5 | 6.2 | 51 | 68.6 | 6.5 | 24.5 | 3.6 |
| Rabies | 4 | 75.0 | 21.7 | 0 | - | - | - | - | 0 | - | - | 0.0 | 0.0 |
| Respiratory Infections | 841 | 64.2 | 1.7 | 476 | 70.0 | 2.1 | 22.9 | 2.2 | 222 | 49.1 | 3.4 | 16.4 | 1.4 |
| Rotavirus | 2 | 100.0 | 0.0 | 2 | 0.0 | 0.0 | 0.0 | 0.0 | 0 | - | - | 0.0 | 0.0 |
| Sepsis | 334 | 66.8 | 2.6 | 206 | 64.6 | 3.3 | 10.2 | 2.4 | 81 | 25.9 | 4.9 | 7.9 | 1.7 |
| Severe Acute Respiratory Syndrome (SARS) | 1 | 100.0 | 0.0 | 1 | 0.0 | 0.0 | 0.0 | 0.0 | 0 | - | - | 0.0 | 0.0 |
| Smallpox | 2 | 100.0 | 0.0 | 2 | 100.0 | 0.0 | 100.0 | 0.0 | 2 | 100.0 | 0.0 | 100.0 | 0.0 |
| Urinary Tract Infections | 276 | 55.1 | 3.0 | 143 | 72.0 | 3.8 | 10.5 | 3.2 | 51 | 29.4 | 6.4 | 7.0 | 1.7 |
| West Nile Virus (WNV) | 2 | 50.0 | 35.4 | 1 | 0.0 | 0.0 | 0.0 | 0.0 | 0 | - | - | 0.0 | 0.0 |
| Yellow Fever | 2 | 0.0 | 0.0 | 0 | - | - | - | - | 0 | - | - | 0.0 | 0.0 |
| Total | 3,851 | 65.0 | 0.8 | 2,202 | 64.3 | 1.0 | 23.2 | 1.0 | 998 | 51.1 | 1.6 | 16.3 | 0.7 |

**Table 7.**
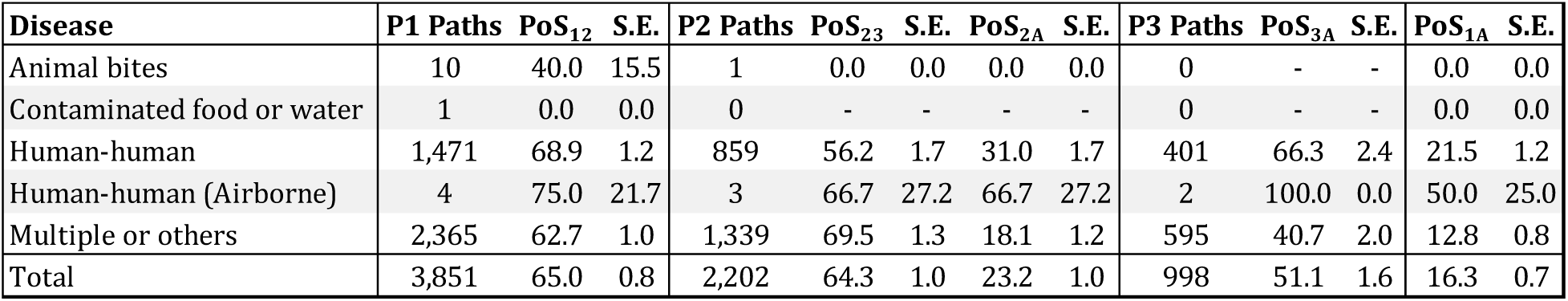
The probabilities of success (PoS) of industry-sponsored, non-vaccine anti-infective drug development programs, grouped by the transmission route. A: regulatory approval; P1: phase 1; P2: phase 2; P3: phase 3; SE: standard error.

| Disease | P1 Paths | PoS<sub>12</sub> | S.E. | P2 Paths | PoS<sub>23</sub> | S.E. | PoS<sub>2A</sub> | S.E. | P3 Paths | PoS<sub>3A</sub> | S.E. | PoS<sub>1A</sub> | S.E. |
| --- | --- | --- | --- | --- | --- | --- | --- | --- | --- | --- | --- | --- | --- |
| Animal bites | 10 | 40.0 | 15.5 | 1 | 0.0 | 0.0 | 0.0 | 0.0 | 0 | - | - | 0.0 | 0.0 |
| Contaminated food or water | 1 | 0.0 | 0.0 | 0 | - | - | - | - | 0 | - | - | 0.0 | 0.0 |
| Human-human | 1,471 | 68.9 | 1.2 | 859 | 56.2 | 1.7 | 31.0 | 1.7 | 401 | 66.3 | 2.4 | 21.5 | 1.2 |
| Human-human (Airborne) | 4 | 75.0 | 21.7 | 3 | 66.7 | 27.2 | 66.7 | 27.2 | 2 | 100.0 | 0.0 | 50.0 | 25.0 |
| Multiple or others | 2,365 | 62.7 | 1.0 | 1,339 | 69.5 | 1.3 | 18.1 | 1.2 | 595 | 40.7 | 2.0 | 12.8 | 0.8 |
| Total | 3,851 | 65.0 | 0.8 | 2,202 | 64.3 | 1.0 | 23.2 | 1.0 | 998 | 51.1 | 1.6 | 16.3 | 0.7 |

**Table 8.** The probabilities of success (PoS) of industry-sponsored, non-vaccine anti-infective drug development programs, grouped by the biological family. A: regulatory approval; P1: phase 1; P2: phase 2; P3: phase 3; SE: standard error.

| Disease | P1 Paths | PoS<sub>12</sub> | S.E. | P2 Paths | PoS<sub>23</sub> | S.E. | PoS<sub>2A</sub> | S.E. | P3 Paths | PoS<sub>3A</sub> | S.E. | PoS<sub>1A</sub> | S.E. |
| --- | --- | --- | --- | --- | --- | --- | --- | --- | --- | --- | --- | --- | --- |
| Caliciviridae | 1 | 0.0 | 0.0 | 0 | - | - | - | - | 0 | - | - | 0.0 | 0.0 |
| Clostridiaceae | 91 | 83.5 | 3.9 | 66 | 53.0 | 6.1 | 4.5 | 2.8 | 25 | 12.0 | 6.5 | 4.2 | 2.4 |
| Coronaviridae | 2 | 50.0 | 35.4 | 1 | 0.0 | 0.0 | 0.0 | 0.0 | 0 | - | - | 0.0 | 0.0 |
| Filoviridae | 10 | 20.0 | 12.6 | 1 | 0.0 | 0.0 | 0.0 | 0.0 | 0 | - | - | 0.0 | 0.0 |
| Flaviviridae | 609 | 70.3 | 1.9 | 383 | 49.6 | 2.6 | 22.5 | 2.2 | 161 | 53.4 | 3.9 | 16.1 | 1.6 |
| Hepadnaviridae | 186 | 77.4 | 3.1 | 105 | 68.6 | 4.5 | 36.2 | 5.2 | 54 | 70.4 | 6.2 | 29.5 | 4.0 |
| Herpesviridae | 64 | 87.5 | 4.1 | 43 | 60.5 | 7.5 | 32.6 | 7.8 | 19 | 73.7 | 10.1 | 31.8 | 7.0 |
| Poxviridae | 3 | 66.7 | 27.2 | 2 | 100.0 | 0.0 | 100.0 | 0.0 | 2 | 100.0 | 0.0 | 66.7 | 27.2 |
| Reoviridae | 2 | 100.0 | 0.0 | 2 | 0.0 | 0.0 | 0.0 | 0.0 | 0 | - | - | 0.0 | 0.0 |
| Retroviridae | 604 | 63.2 | 2.0 | 326 | 59.8 | 2.7 | 39.3 | 2.8 | 167 | 76.6 | 3.3 | 24.6 | 1.9 |
| Rhabdoviridae | 4 | 75.0 | 21.7 | 0 | - | - | - | - | 0 | - | - | 0.0 | 0.0 |
| Togaviridae | 1 | 0.0 | 0.0 | 0 | - | - | - | - | 0 | - | - | 0.0 | 0.0 |
| Multiple or others | 2,274 | 61.9 | 1.0 | 1,273 | 70.3 | 1.3 | 18.8 | 1.3 | 570 | 41.9 | 2.1 | 13.2 | 0.8 |
| Total | 3,851 | 65.0 | 0.8 | 2,202 | 64.3 | 1.0 | 23.2 | 1.0 | 998 | 51.1 | 1.6 | 16.3 | 0.7 |

### A5. PoS Tables for vaccines (non-industry-sponsored)

**Table 9.** The probabilities of success (PoS) of non-industry-sponsored vaccine development programs. A: regulatory approval; P1: phase 1; P2: phase 2; P3: phase 3; SE: standard error.

| Disease | P1 Paths | PoS<sub>12</sub> | S.E. | P2 Paths | PoS<sub>23</sub> | S.E. | PoS<sub>2A</sub> | S.E. | P3 Paths | PoS<sub>3A</sub> | S.E. | PoS<sub>1A</sub> | S.E. |
| --- | --- | --- | --- | --- | --- | --- | --- | --- | --- | --- | --- | --- | --- |
| Bacterial Skin Infection | 3 | 100.0 | 0.0 | 0 | - | - | - | - | 0 | - | - | 0.0 | 0.0 |
| Clostridium difficile | 3 | 66.7 | 27.2 | 0 | - | - | - | - | 0 | - | - | 0.0 | 0.0 |
| Cytomegalovirus Infection (CMV) | 14 | 50.0 | 13.4 | 5 | 40.0 | 21.9 | 40.0 | 21.9 | 2 | 100.0 | 0.0 | 16.7 | 10.8 |
| Ebola | 8 | 12.5 | 11.7 | 0 | - | - | - | - | 0 | - | - | 0.0 | 0.0 |
| Hepatitis B Virus (HBV) | 58 | 91.4 | 3.7 | 48 | 47.9 | 7.2 | 8.3 | 4.3 | 16 | 25.0 | 10.8 | 8.7 | 4.2 |
| Hepatitis C Virus (HCV) | 10 | 70.0 | 14.5 | 5 | 0.0 | 0.0 | 0.0 | 0.0 | 0 | - | - | 0.0 | 0.0 |
| HIV | 144 | 48.6 | 4.2 | 62 | 3.2 | 2.2 | 0.0 | 0.0 | 2 | 0.0 | 0.0 | 0.0 | 0.0 |
| Human Papillomavirus (HPV) | 32 | 87.5 | 5.8 | 16 | 56.3 | 12.4 | 6.3 | 6.5 | 7 | 14.3 | 13.2 | 5.6 | 5.4 |
| Intra-abdominal Infections | 3 | 100.0 | 0.0 | 0 | - | - | - | - | 0 | - | - | 0.0 | 0.0 |
| Japanese Encephalitis | 4 | 100.0 | 0.0 | 4 | 100.0 | 0.0 | 25.0 | 21.7 | 4 | 25.0 | 21.7 | 25.0 | 21.7 |
| Middle East Respiratory Syndrome (MERS) | 1 | 100.0 | 0.0 | 0 | - | - | - | - | 0 | - | - | 0.0 | 0.0 |
| Otitis Media | 7 | 100.0 | 0.0 | 7 | 28.6 | 17.1 | 28.6 | 17.1 | 2 | 100.0 | 0.0 | 28.6 | 17.1 |
| Rabies | 16 | 81.3 | 9.8 | 13 | 53.8 | 13.8 | 30.8 | 12.8 | 7 | 57.1 | 18.7 | 25.0 | 10.8 |
| Respiratory Infections | 175 | 66.9 | 3.6 | 101 | 51.5 | 5.0 | 16.8 | 3.9 | 41 | 41.5 | 7.7 | 11.5 | 2.6 |
| Rotavirus | 5 | 80.0 | 17.9 | 4 | 50.0 | 25.0 | 0.0 | 0.0 | 1 | 0.0 | 0.0 | 0.0 | 0.0 |
| Sepsis | 7 | 42.9 | 18.7 | 2 | 0.0 | 0.0 | 0.0 | 0.0 | 0 | - | - | 0.0 | 0.0 |
| Severe Acute Respiratory Syndrome (SARS) | 1 | 0.0 | 0.0 | 0 | - | - | - | - | 0 | - | - | 0.0 | 0.0 |
| Smallpox | 11 | 63.6 | 14.5 | 6 | 16.7 | 15.2 | 16.7 | 15.2 | 1 | 100.0 | 0.0 | 10.0 | 9.5 |
| Urinary Tract Infections | 3 | 100.0 | 0.0 | 1 | 0.0 | 0.0 | 0.0 | 0.0 | 0 | - | - | 0.0 | 0.0 |
| West Nile Virus (WNV) | 4 | 0.0 | 0.0 | 0 | - | - | - | - | 0 | - | - | 0.0 | 0.0 |
| Yellow Fever | 9 | 66.7 | 15.7 | 6 | 33.3 | 19.2 | 0.0 | 0.0 | 1 | 0.0 | 0.0 | 0.0 | 0.0 |
| Zika | 5 | 40.0 | 21.9 | 0 | - | - | - | - | 0 | - | - | 0.0 | 0.0 |
| Others | 183 | 57.9 | 3.6 | 71 | 35.2 | 5.7 | 9.9 | 3.8 | 14 | 50.0 | 13.4 | 5.1 | 1.9 |
| Total | 706 | 63.3 | 1.8 | 351 | 37.3 | 2.6 | 11.1 | 1.8 | 98 | 39.8 | 4.9 | 6.8 | 1.0 |

### A6. PoS Tables for the treatment of infectious diseases (non-industry-sponsored)

**Table 10.** The probabilities of success (PoS) of non-industry-sponsored, non-vaccine anti-infective drug development programs. A: regulatory approval; P1: phase 1; P2: phase 2; P3: phase 3; SE: standard error.

| Disease | P1 Paths | PoS<sub>12</sub> | S.E. | P2 Paths | PoS<sub>23</sub> | S.E. | PoS<sub>2A</sub> | S.E. | P3 Paths | PoS<sub>3A</sub> | S.E. | PoS<sub>1A</sub> | S.E. |
| --- | --- | --- | --- | --- | --- | --- | --- | --- | --- | --- | --- | --- | --- |
| Bacterial Skin Infection | 360 | 46.4 | 2.6 | 151 | 81.5 | 3.2 | 19.2 | 3.7 | 85 | 34.1 | 5.1 | 9.5 | 1.7 |
| Chikungunya | 2 | 100.0 | 0.0 | 2 | 50.0 | 35.4 | 0.0 | 0.0 | 1 | 0.0 | 0.0 | 0.0 | 0.0 |
| Clostridium difficile | 83 | 94.0 | 2.6 | 51 | 76.5 | 5.9 | 15.7 | 6.2 | 22 | 36.4 | 10.3 | 20.5 | 6.5 |
| Cytomegalovirus Infection (CMV) | 77 | 83.1 | 4.3 | 51 | 51.0 | 7.0 | 23.5 | 6.9 | 13 | 92.3 | 7.4 | 23.5 | 5.9 |
| Ebola | 30 | 96.7 | 3.3 | 28 | 14.3 | 6.6 | 0.0 | 0.0 | 2 | 0.0 | 0.0 | 0.0 | 0.0 |
| Hepatitis B Virus (HBV) | 171 | 49.1 | 3.8 | 73 | 47.9 | 5.8 | 1.4 | 1.4 | 31 | 3.2 | 3.2 | 0.6 | 0.6 |
| Hepatitis C Virus (HCV) | 139 | 84.2 | 3.1 | 112 | 43.8 | 4.7 | 8.9 | 2.9 | 33 | 30.3 | 8.0 | 8.5 | 2.6 |
| Human Immunodeficiency Virus (HIV) | 414 | 61.1 | 2.4 | 195 | 49.2 | 3.6 | 13.3 | 2.6 | 75 | 34.7 | 5.5 | 7.8 | 1.5 |
| Human Papillomavirus (HPV) | 71 | 88.7 | 3.8 | 42 | 42.9 | 7.6 | 2.4 | 2.7 | 8 | 12.5 | 11.7 | 2.5 | 2.5 |
| Intra-abdominal Infections | 189 | 66.1 | 3.4 | 112 | 76.8 | 4.0 | 12.5 | 3.8 | 51 | 27.5 | 6.2 | 9.9 | 2.5 |
| Japanese Encephalitis | 2 | 100.0 | 0.0 | 2 | 0.0 | 0.0 | 0.0 | 0.0 | 0 | - | - | 0.0 | 0.0 |
| Middle East Respiratory Syndrome (MERS) | 3 | 100.0 | 0.0 | 3 | 66.7 | 27.2 | 0.0 | 0.0 | 0 | - | - | 0.0 | 0.0 |
| Non-tuberculous Mycobacteria (NTM) Infection | 13 | 84.6 | 10.0 | 9 | 44.4 | 16.6 | 11.1 | 11.9 | 2 | 50.0 | 35.4 | 11.1 | 10.5 |
| Norovirus | 2 | 100.0 | 0.0 | 1 | 0.0 | 0.0 | 0.0 | 0.0 | 0 | - | - | 0.0 | 0.0 |
| Onychomycosis | 20 | 75.0 | 9.7 | 15 | 66.7 | 12.2 | 0.0 | 0.0 | 6 | 0.0 | 0.0 | 0.0 | 0.0 |
| Otitis Media | 186 | 30.1 | 3.4 | 53 | 56.6 | 6.8 | 7.5 | 3.9 | 24 | 16.7 | 7.6 | 2.3 | 1.1 |
| Rabies | 1 | 0.0 | 0.0 | 0 | - | - | - | - | 0 | - | - | 0.0 | 0.0 |
| Respiratory Infections | 610 | 58.5 | 2.0 | 323 | 72.8 | 2.5 | 11.5 | 2.1 | 141 | 26.2 | 3.7 | 7.7 | 1.2 |
| Sepsis | 310 | 80.0 | 2.3 | 227 | 77.5 | 2.8 | 15.9 | 3.0 | 94 | 38.3 | 5.0 | 17.4 | 2.6 |
| Severe Acute Respiratory Syndrome (SARS) | 3 | 100.0 | 0.0 | 3 | 100.0 | 0.0 | 0.0 | 0.0 | 2 | 0.0 | 0.0 | 0.0 | 0.0 |
| Urinary Tract Infections | 291 | 46.7 | 2.9 | 126 | 73.8 | 3.9 | 10.3 | 3.4 | 49 | 26.5 | 6.3 | 5.5 | 1.5 |
| West Nile Virus (WNV) | 1 | 100.0 | 0.0 | 1 | 0.0 | 0.0 | 0.0 | 0.0 | 0 | - | - | 0.0 | 0.0 |
| Total | 2,978 | 61.0 | 0.9 | 1,580 | 65.2 | 1.2 | 12.2 | 0.9 | 639 | 30.0 | 1.8 | 8.2 | 0.6 |

### A7. Regression of the difference between the number of industry-sponsored investigational treatments initiated and industry-sponsored investigational vaccines initiated against time

We attempt to find correlations between the difference in the number of industry-sponsored clinical development programs initiated for treatments and the analogous number for vaccines (*DIFF*). We define ‘*TIME*’ as the number of months elapsed since January 1, 2000.

We test five regression models with the follow specifications:

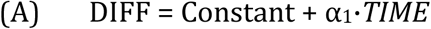

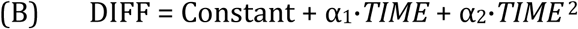

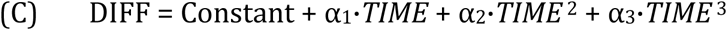

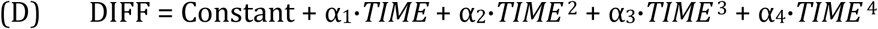

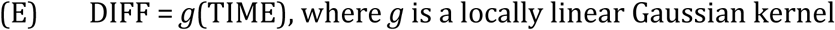

Models (A) to (D) are performed on Microsoft Excel 2019 while Model (E) is implemented using the statsmodel package in Python. The bandwidth for the non-parametric kernel regression is selected using the least-squares cross-validation method. The results of the regressions are reported in Table 11. Our best fitting model, Model E, indicates that the difference between the number of non-vaccine treatment development programs initiated and the number of vaccine programs initiated widened between January 2000 and May 2014 before narrowing. The difference between the number of initiated programs is close to zero in December 2019, possibly due to the withdrawal of pharmaceutical companies from the antibiotics business and bankruptcy of antibiotics companies.

**Table 11.** Regression results for the five models specified. The standard errors of the coefficients are enclosed in parentheses. An asterisk indicates that the coefficient is statistically significant at the 5% confidence level.

| Regression Statistics | Model A | Model B | Model C | Model D | Model E |
| --- | --- | --- | --- | --- | --- |
| R-Squared | 0.067 | 0.090 | 0.146 | 0.166 | 0.178 |
| Adjusted R-Square | 0.063 | 0.083 | 0.135 | 0.151 | 0.175 |
| Standard Error | 8.75 | 8.65 | 8.40 | 8.32 | 8.22 |
| Coefficients |  |  |  |  |  |
| Intercept | *3.58<br>(1.13) | 0.52<br>(1.66) | *6.00<br>(2.14) | 2.34<br>(2.62) |  |
| Time | *3.36E-02<br>(8.15E-03) | *1.11E-01<br>(3.21E-02) | *-1.67E-01<br>(7.76E-02) | 1.45E-01<br>(1.53E-01) |  |
| Time<sup>2</sup> |  | *-3.23E-04<br>(1.30E-04) | *2.59E-03<br>(7.55E-04) | -3.31E-03<br>(2.60E-03) |  |
| Time<sup>3</sup> |  |  | *-8.12E-06<br>(2.08E-06) | 3.03E-05<br>(1.64E-05) |  |
| Time<sup>4</sup> |  |  |  | *-8.05E-08<br>(3.40E-08) |  |

